# Copy number variant association analysis in 94,730 Chinese adults reveals loci influencing anthropometric and cardiometabolic traits

**DOI:** 10.64898/2026.08.12.26359684

**Authors:** Isobel Howard, Iona Y. Millwood, Sam Morris, Kuang Lin, Daniel Avery, Canqing Yu, Jun Lv, Dianjianyi Sun, Pei Pei, Liming Li, Junshi Chen, Zhengming Chen, Robin G. Walters, Fiona Bragg, Derrick Bennett

## Abstract

Copy-number variants (CNVs) represent an important source of genetic variation that can influence complex traits and disease risk by altering gene dosage, disrupting coding sequence, or modifying regulatory elements. Existing CNV association studies have been limited in scale and have largely focused on European-ancestry populations. We present a CNV genome-wide association study of 13 anthropometric and cardiometabolic traits in 94,730 adults from the China Kadoorie Biobank, a large East Asian study. We identify 19 independent locus-phenotype associations across 15 unique loci. Novel associations include random plasma glucose at 8p23.1 (β = -0.29 SD, *P* = 5.40×10⁻) and 14q11.2 (β = +0.43 SD, *P* = 8.41×10⁻), diastolic blood pressure at 7p21.1 (β = +0.75 SD, *P* = 5.25×10⁻), and duplication-associated reductions in body fat percentage at 12p12.1 (β = -0.74 SD, *P* = 8.11×10⁻) and 17q12 (β = -0.56 SD, *P* = 7.36×10⁻). We also replicated established dosage-sensitive regions, most prominently at two distinct intervals within 16p11.2 (BP2-BP3 and BP4-BP5), where CNVs show large bidirectional dosage effects across 5 adiposity traits including body mass index (β = -0.84 SD per copy, *P* = 1.77×10⁻). These findings identify structural variants contributing to cardiometabolic and anthropometric trait variation in Chinese adults and expand the ancestry diversity of CNV association studies.

## Introduction

Genome-wide association studies (GWAS) have established the highly polygenic architecture of complex human traits, yet common single-nucleotide polymorphisms (SNPs) explain only part of phenotypic variance^1–3^. Copy-number variants (CNVs), defined as deletions or duplications of genomic segments typically ≥ 1000 base pairs in size, represent an important additional source of genetic variation. By altering gene dosage, disrupting coding sequence, or modifying regulatory elements, CNVs can have substantial functional consequences^4–6^. Moreover, since they often span many kilobases to megabases, they can have larger functional effects than SNPs and have been implicated in a wide range of complex traits and diseases, suggesting that they capture phenotypic variation not explained by common SNPs alone^7,8^.

Historically, many studies have focused on well-characterised recurrent or pathogenic CNVs, first identified through their large effects on neurodevelopmental and psychiatric disorders^9–12^. These CNVs have since been implicated in a broader range of phenotypes, including cardiometabolic traits^9,10^. The expansion of large population biobanks and consortia of these has enabled genome-wide analyses, allowing more comprehensive assessment of CNV-trait associations without prior locus selection^13–18^. However, these large-scale CNV association studies remain heavily concentrated in European-ancestry populations, and studies in East Asian populations have largely been limited in scope. For example, population-wide CNV mapping efforts in East Asian populations have examined samples of fewer than 10,000 individuals^19–23^, which are insufficient for comprehensive metabolic trait analysis or reliable detection of rare CNVs with moderate effect sizes. Disease-specific studies in East Asian populations have similarly been constrained by investigation in small case-control studies^24–26^, precluding the phenome-wide discovery approaches that have been implemented in European cohorts. This represents a meaningful gap, given SNP-based GWAS demonstrates that allele frequencies, effect sizes, and locus architectures can differ substantially across ancestries^27,28^. CNV frequencies have similarly been shown to vary across populations, suggesting that CNV-trait associations identified in European cohorts may not be transferable^29,30^.

Here, we address two aims in 94,793 genotyped adults from the China Kadoorie Biobank (CKB): (1) to characterise the genome-wide CNV landscape in a large East Asian population; and (2) to identify CNV associations with 13 anthropometric and cardiometabolic traits; thereby expanding the ancestry diversity of CNV association studies beyond the predominantly European-ancestry cohorts studied to date.

## Methods

### Study Material

#### Cohort Description

CKB is a prospective cohort of 512,714 individuals aged 30-79 years recruited from 10 geographically diverse regions of China between 2004 and 2008^31,32^. At baseline, participants completed an interviewer-administered questionnaire covering sociodemographic factors, lifestyle, and medical history, and underwent physical examination with collection of blood samples for long-term storage. All participants provided written informed consent. This study was approved by the Oxford Tropical Research Ethics Committee, Oxford University (UK, 025-04) and the China National Center for Disease Control and Prevention (CDC) (Beijing, China, 005/2004). All analyses used data derived from CKB Data Release 19.01.

#### Phenotype Measurements

All physical measurements were obtained by trained health workers at local study assessment clinics using calibrated instruments and standardised protocols, as described previously^31,32^. For the present analyses, we selected 13 quantitative traits from baseline measurements, encompassing anthropometric and body composition measures (body mass index [BMI], BMI at age 25 years, standing height, sitting height, weight, waist circumference, hip circumference, waist-hip ratio [WHR], and body fat percentage), cardiovascular measures (systolic blood pressure [SBP], diastolic blood pressure [DBP], and resting heart rate), and a glycaemic measure (random plasma glucose [RPG]).

Anthropometric and body composition measurementswere undertaken with participants wearing light clothing and no shoes. Standing height, sitting height, and body weight were measured using standardised instruments. Waist circumference was measured to the nearest 0.1 cm with a non-stretchable tape measure at the midpoint between the lowest rib margin and the iliac crest; hip circumference was measured similarly around the maximum circumference of the buttocks. Body fat percentage was estimated using foot-to-foot bioelectrical impedance analysis (BIA; TBF-300 Body Composition Analyzer, Tanita Inc., Tokyo, Japan). BMI was calculated as weight (kg) divided by height squared (m²). BMI at age 25 years was derived from self-reported recalled weight at age 25 and measured height at baseline. WHR was calculated as waist circumference divided by hip circumference.

Blood pressure and resting heart rate were measured using an Omron UA-779 digital sphygmomanometer after participants had been seated at rest for at least five minutes. Two measurements were taken; if the difference between the two SBP readings exceeded 10 mmHg, a third measurement was obtained. The mean of the last two recorded values was used in all analyses.

A 10 mL non-fasting venous blood sample was collected at baseline, with time since last food intake recorded. RPG was measured on-site using a SureStep Plus meter (LifeScan, Johnson & Johnson), regularly calibrated with manufacturer control solutions.

#### Genotyping Data

Genotyping procedures and quality control (QC) have been described in detail previously^33^. Briefly, participants were genotyped using 2 custom-designed Affymetrix Axiom arrays, optimised for individuals of East Asian ancestry: CKB1, used primarily in nested case-control studies and enriched for individuals with major cardiovascular and respiratory disease (33,408 participants), and CKB2, with improved probe density, applied predominantly to population-representative samples (72,000 participants). Raw CEL files were processed using Axiom Analysis Suite^34^, which applies background correction, plate-level normalisation, and marker-wise scaling before fitting allele-specific intensity clusters using the Axiom GT1 algorithm. The output signal intensity data for each sample were processed using Axiom CNV Tools v1.1 to derive normalised log R ratio (LRR) and B-allele frequency (BAF) values for each probe, reflecting total signal intensity and allelic balance respectively. At the sample level, arrays failing manufacturer thresholds for DishQC (< 0.82) or call rate (< 97%) were excluded. At the probe level, markers with poor clustering performance, batch-specific artefacts, or missingness exceeding 5% were removed. Samples identified as duplicates, sex mismatches, or exhibiting excess heterozygosity, homozygosity, or chromosomal aneuploidy were further excluded^33^. This upstream quality control pipeline yielded normalised LRR and BAF signal files for 100,706 individuals (32,300 on CKB1 and 68,406 on CKB2), which served as input to the CNV calling pipeline described below (**Figure S1**). To account for potential technical variability between array versions and recruitment regions, CNV calling was performed separately for each region-array combination, yielding 20 strata analysed independently. All genomic coordinates refer to GRCh37/hg19.

#### CNV Calling and Quality Control

Autosomal CNVs were detected using PennCNV v1.0.5 with PennCNV-Affy^35^, a hidden Markov model-based algorithm that infers copy-number state from normalised LRR and BAF derived from SNP array intensity data as above. This has been shown to outperform alternative callers in precision and overall accuracy in benchmarking studies^36,37^. For each stratum, CNV calling incorporated normalised LRR and BAF signal files, a population frequency of B-allele (PFB) file, a GC-model file to correct for GC-wave artefacts, and the affygw6.hmm hidden Markov model optimised for Affymetrix arrays^35^. The PFB file was constructed from 1,000 randomly selected unrelated individuals within the stratum who passed PennCNV signal quality thresholds^35^.

Following CNV detection, post-calling quality control was applied within each stratum. CNVs overlapping genomic regions prone to technical artefact were excluded: specifically, centromeric regions, telomeric regions (defined as the first and last 500 kb of each chromosome), and segmental duplication hotspots (**Table ST1**). CNVs were excluded if 50% or more of their base pair range overlapped a segmental duplication region, following the approach recommended for Affymetrix arrays^35^. Samples were removed if they met any of the following criteria: LRR standard deviation > 0.35; BAF drift > 0.01; absolute waviness factor > 0.05; or more than 100 detected CNVs (**Figure S2**). After sample-level quality control, 94,793 individuals passed CNV quality control filters; of these, 94,730 had phenotypic data available for at least one trait of interest and contributed to association analyses. CNVs were retained if they had a PennCNV confidence score ≥ 20, spanned at least 10 kb, and were supported by at least 10 probes.

Adjacent CNVs of the same type within a sample were merged prior to quality filtering if the intervening gap was ≤20% of the combined CNV span. This merging step reduces fragmentation of large events, a known behaviour of CNV calling on high-density arrays whereby a single CNV may be called as multiple adjacent segments^35^. CNVs meeting all of the above criteria were retained for association analyses and are hereafter referred to as high-confidence CNVs.

### CNV Association Studies

All analyses were performed in R v4.3.1. Gene annotation was conducted using GENCODE v19 (GRCh37/hg19)^38^.

#### Construction of Probe-Level CNV Matrices

High-confidence CNVs were mapped to array probes based on genomic overlap. For each probe-sample pair, an inferred copy-number state was assigned from PennCNV calls: deletions were defined as CN < 2 (CN = 0 or 1), duplications as CN > 2 (CN ≥ 3), and copy-neutral probes as CN = 2. Exact copy-number states were collapsed into deletion and duplication categories to reflect underlying dosage effects.

Three probe-by-sample matrices were constructed: (i) a mirror model matrix coding deletions as −1, duplications as +1, and copy-neutral probes as 0, designed to capture dosage effects acting in opposite directions under a shared linear encoding; (ii) a deletion-only matrix coding deletions as 1 and excluding duplications (set to missing), to isolate deletion-specific effects; and (iii) a duplication-only matrix coding duplications as 1 and excluding deletions (set to missing), to isolate duplication-specific effects. Individuals passing sample-level quality control but without any detected CNVs contributed copy-neutral values across all probes and were retained in the analysis, ensuring that association tests compare CNV carriers against the full set of non-carriers rather than only those with CNVs of the opposite type.

Because the CKB1 and CKB2 arrays differ in probe content and architecture, association analyses were restricted to the 541,024 probes shared between arrays with consistent genomic coordinates, enabling pooled analyses while preserving probe-level comparability. To account for potential technical variability between arrays, array version was included as a covariate in all association models. To confirm that pooling across array versions did not distort inference, we performed stratified validation by array version at all CNV-wide significant loci, including formal CNV-array version interaction tests (**Supplementary Note 1**). Effect estimates were directionally consistent across array versions and no interaction survived Bonferroni correction (**Figures S3-S5**).

Probes were required to have ≥15 carriers to ensure stable regression estimates, as the precision of effect estimates for a rare carrier predictor depends on the absolute number of carriers rather than carrier frequency. This threshold falls within the range used in comparable large-scale CNV association studies^13,16,17^. A fixed carrier-count threshold, rather than a frequency-based one, was applied given that sample size varied modestly across phenotypes (range: 78,603–94,730; **Table ST3**), ensuring a consistent minimum standard for regression stability irrespective of phenotype-specific sample size. For the mirror model, probes were retained if ≥15 carriers of either type were present, reflecting the joint dosage encoding.

#### Association Analyses

All phenotypes were inverse rank-normal transformed based on raw trait values across the phenotype-specific analysis sample prior to association testing, to improve adherence to linear regression assumptions and reduce the influence of outliers^39^. Probe-level association testing was performed on the pooled dataset using linear regression for RINT-transformed continuous traits. Models were adjusted for sex, age, age^2^, study region (categorical), genotyping array version (CKB1 versus CKB2; categorical), and the first 11 genetic principal components derived from genome-wide genotype data in unrelated CKB participants following iterative exclusion of long-range linkage disequilibrium regions, which were identified as informative for CKB population structure using the Bayesian information criterion^33^. Effect estimates are reported on the RINT scale, which has unit variance and can be interpreted as standardised effect sizes^40^.

Primary analyses included all participants. To assess the potential impact of relatedness, analyses were repeated in a subset of individuals unrelated to the third family degree^33^ (n = 71,477-72,183 depending on trait).

#### Multiple Testing Correction

Probe-level CNV matrices exhibit high inter-probe correlation by design, as CNV events spanning multiple probes induce extended blocks of correlated carrier status among neighbouring probes. The naive number of tests therefore substantially overstates the number of independent hypotheses tested. Multiple testing correction was performed using the SimpleM method^41^, which estimates the effective number of independent tests (M_eff_) by eigenvalue decomposition of the inter-probe correlation matrix, retaining the number of principal components required to explain 99.5% of total variance. Chromosome-level M_eff_ estimates were summed across autosomes (chromosomes 1-22). Association testing was performed on 31,923, 46,103, and 80,612 probes passing the minimum carrier threshold under the deletion, duplication, and mirror models, respectively, across the full analysis sample; probe counts varied modestly across phenotypes owing to differences in sample size after missing data exclusion. SimpleM M_eff_ estimation was performed separately for each phenotype-model combination using the probes passing the carrier threshold for that phenotype. The maximum M_eff_ across phenotypes was taken as a conservative, phenotype-invariant threshold. This yielded M_eff_ = 5,954 (deletion), M_eff_ = 3,765 (duplication), and M_eff_ = 5,404 (mirror), giving CNV-wide significance thresholds of *P* ≤ 8.40×10⁻, *P* ≤ 1.33×10⁻, and *P* ≤ 9.24×10⁻, respectively. These CNV-wide thresholds were used as the primary criterion for signal inclusion.

To additionally account for the number of phenotypes examined (n=13), an experiment-wide threshold was derived by applying SimpleM^41^ to the phenotypic correlation matrix, yielding M_eff_ = 11 effective traits from the 13 phenotypes. Each model-specific threshold was divided accordingly. Given the high between-phenotype correlation among the traits examined, this adjustment is conservative and CNV-wide thresholds are used as the primary criterion throughout.

#### CNVR Definition

CNV-wide significant probes were consolidated into copy-number variable regions (CNVRs) using a tagging approach based on linkage disequilibrium (LD) following Auwerx et al.^17^, designed to define regions around independent lead probes rather than relying on probe contiguity.

Within each phenotype and association model (deletion-only, duplication-only, and mirror), CNV-wide significant probes were considered as candidate lead probes and processed chromosome by chromosome. Probes were ranked by association *p*-value and lead probes selected iteratively. At each step, the most significant unassigned probe was designated a lead probe; all probes within ±3 Mbp were evaluated for correlation with the lead probe using the corresponding model-specific CNV carrier matrix, following the approach of Auwerx et al.^17^. Probes with r² ≥ 0.5 were assigned to the same CNVR and excluded from subsequent iterations, continuing until all significant probes were assigned. CNVR boundaries were then defined by extending each lead probe to encompass all probes in the full 541,024-probe shared matrix within ±3 Mbp with r² ≥ 0.5 to the lead^17^. CNVRs were consolidated across models at the phenotype level, merging signals involving the same trait, overlapping CNVRs, and directionally concordant effects under the mirror encoding.

#### Overlap with Known SNP-Trait Associations

To contextualise CNV association signals with prior SNP-based GWAS findings, trait-matched SNP associations were identified from the National Human Genome Research Institute-European Bioinformatics Institute (NHGRI-EBI GWAS Catalog)^42^ (hg19/GRCh37 coordinates, downloaded April 2025) within the r^2^-defined CNVR interval for each locus-phenotype association. Catalog entries were filtered to traits matching a predefined synonym list for each analysed phenotype (**Table ST2**). This analysis was conducted for contextualisation purposes; formal statistical colocalisation was not performed.

## Results

### CNV Landscape

After sample-and call-level quality control, 94,793 participants remained for analysis (**Figures S1, S2**). The analysis population had a mean age of 53.7 years (SD 11.0) and was 57.2% female, recruited across 10 regions of China (5 urban, 5 rural) (**Table ST3**). Of these 94,793 participants, 84,441 carried at least one high-confidence CNV (277,296 CNVs in total), while 10,352 individuals had no CNVs detected. Heterozygous deletions and duplications accounted for the majority of calls (59.7% and 37.7%, respectively), whereas homozygous events were rare (2.6% combined) (**Figure 1A**). As homozygous CNV calls (copy-number states 0 or 4) comprised only 2.6% of the total, we define deletions as CN < 2 and duplications as CN > 2 throughout. Participants carried a mean of 2.93 CNVs per individual (median 3) (**Figure 1B**). Amongst identified high-confidence CNVs, duplications were substantially longer than deletions (median 210 kb versus 77 kb), accounting for 58.9% of the total genomic burden despite representing only 36.4% of calls (**Figure 1C**). CNV carriers had a mean genomic burden of 677 kb (median 415 kb) of affected sequence (**Figure 1D**).

**Figure 1.**
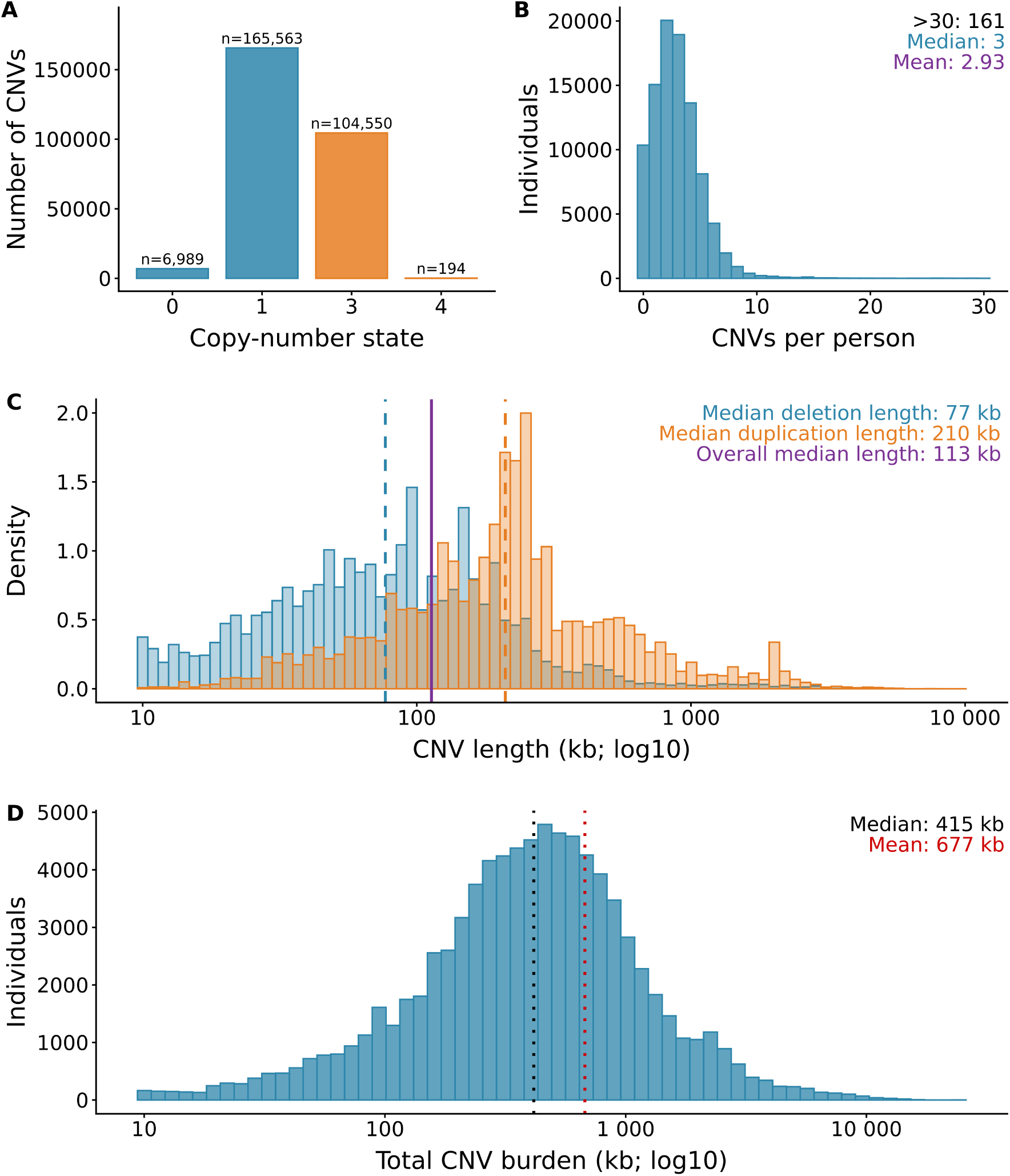
CNV callset characteristics in the CKB cohort. CNV call characteristics across 94,793 individuals and 277,296 CNVs. (A) Distribution of CNV copy-number states. (B) CNV count per individual. (C) CNV length distribution by event type. (D) Total CNV burden per individual.

To assess CNV recurrence across the genome, we projected all calls onto the genotyping-array probe backbone. Probe coverage differed between the two arrays (598,873 and 797,915 probes, with 541,024 shared), but per-probe CNV frequency distributions were highly comparable across array versions. Across the full probe sets, mean deletion frequencies were 0.00012 for both array versions, and mean duplication frequencies were 0.00018 and 0.00012 for CKB1 and CKB2, respectively, with median values close to zero in all cases. Maximum per-probe frequencies reached 0.13 for deletions and 0.16 for duplications, corresponding to recurrent hotspots in regions such as 6p21.3 and 14q11.2 (**Figure 2**). When restricted to the 541,024 shared probes, combined mean frequencies were 0.00010 for deletions and 0.00012 for duplications, with maxima of 0.05 and 0.16, respectively. Only 645 probes (0.12%) had deletion frequencies exceeding 1%, compared with 1,042 probes (0.19%) for duplications.

**Figure 2.**
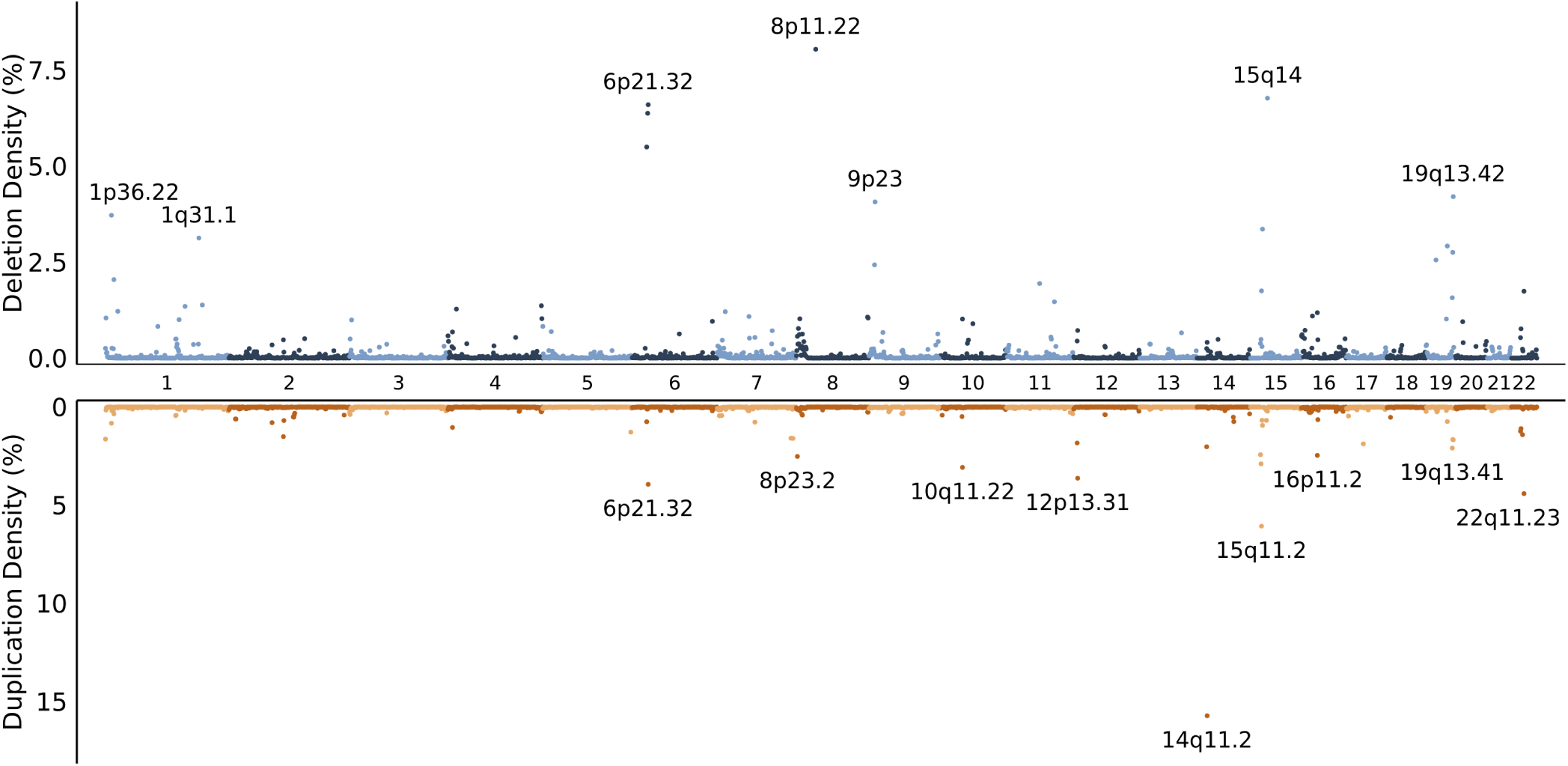
Genome-wide CNV density in 1 Mb bins. Genome-wide CNV density across the CKB cohort, calculated in 1 Mb genomic bins as the percentage of individuals with a CNV overlapping each bin. Upper panel: deletions. Lower panel: duplications, with the y-axis reversed.

CNVs were distributed pervasively across the genome, with elevated densities at a small number of well-established hotspot regions but with measurable burden present on every chromosome (**Figure 2**). Prominent CNV frequency peaks were observed at well-established dosage-sensitive loci including 16p11.2, 22q11.2, and 15q11.2-15q14^43–45^. Additional recurrent regions included 8p23.1-8p23.2, which harbours defensin and olfactory receptor gene clusters within a structurally complex region flanked by segmental duplications and containing a well-documented common chromosomal inversion polymorphism, predisposing to recurrent rearrangement^46,47^. A dense CNV cluster was also detected at 6p21.3 within the major histocompatibility complex (MHC), consistent with known structural diversity in immune-related genes^48^. Subtelomeric regions such as 1p36 and 19q13 likewise showed elevated CNV densities (**Figure 2**). Across most loci, duplication frequencies exceeded deletion frequencies, and recurrent regions typically showed enrichment for one CNV type rather than balanced deletion-duplication frequencies^49^. Together, these patterns recapitulate established CNV hotspots while defining the structural variation landscape of the CKB cohort.

#### Probe-level Association Analysis Overview

Of the 94,793 individuals passing CNV quality control, 94,730 had phenotypic data available for at least one analysed trait; sample sizes across traits ranged from 78,603 (BMI at age 25, reflecting missing recalled weight data) to 94,730, (**Table ST3**). Across all traits, 26 model-specific CNVR-phenotype associations exceeded the corresponding CNV-wide significance threshold, resolving to 19 unique locus-phenotype associations after consolidation of signals detected under both a single-type and mirror model at the same locus (**Figure 3**; **Table 1**). Of the 19 associations, 4 were detected under the deletion model only, 3 under the duplication model only, and 5 under the mirror model only; the remaining 7 were concordantly significant under both a single-type and mirror model (5 duplication+mirror, 2 deletion+mirror). No association was detected across all 3 models simultaneously. Moreover, 6 associations passed the more stringent study-wide threshold further correcting for the number of traits examined (0.05/[M^eff^×11]); given the high inter-phenotype correlation between the 13 traits analysed, this threshold is conservative, and the model-specific CNV-wide thresholds serve as the primary criterion for signal inclusion throughout. Full results for all single CNV type and mirror model associations are provided in **Table ST4** and **Table ST5**, respectively.

**Figure 3.**
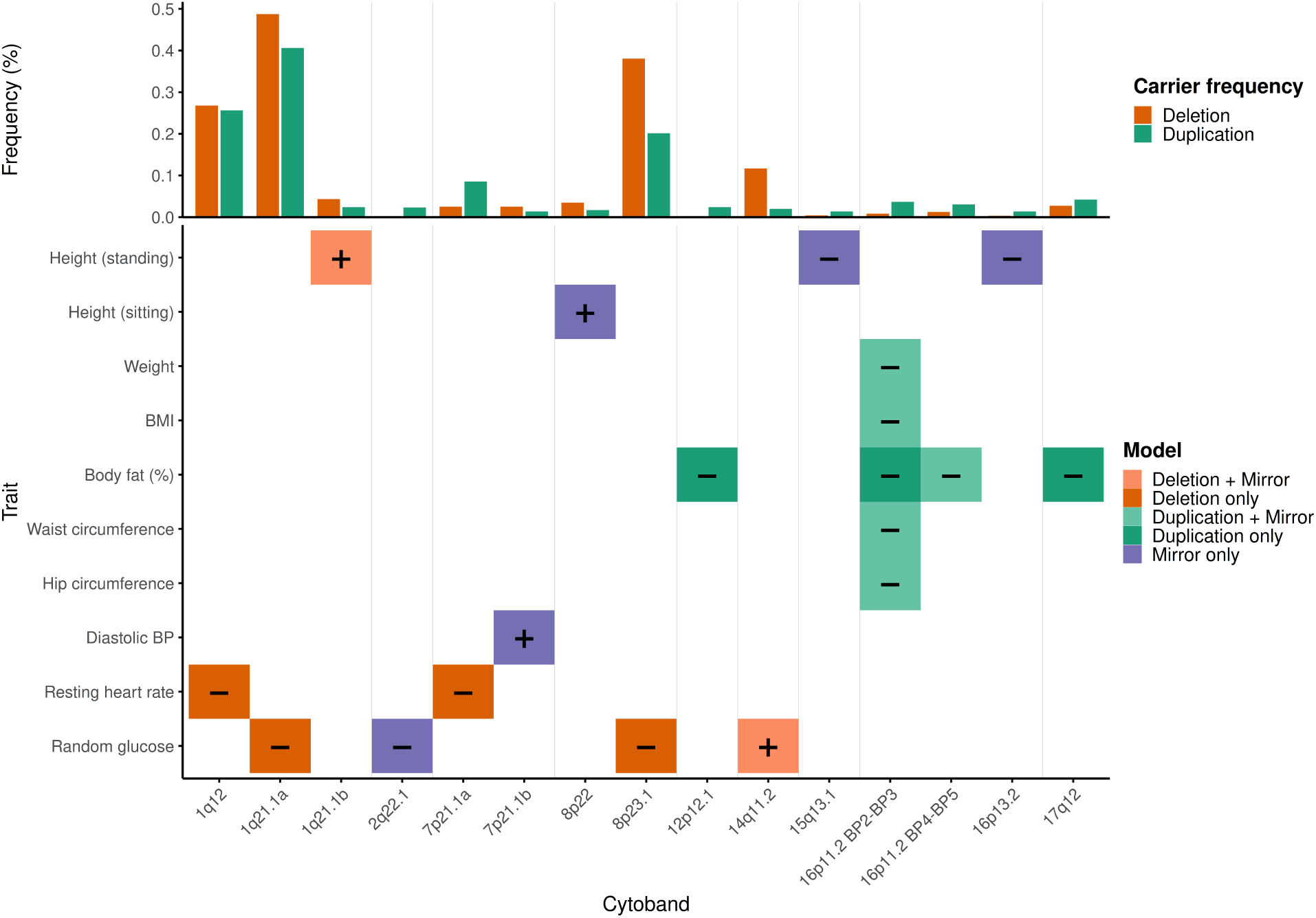
Genome-wide CNVR-trait associations in the China Kadoorie Biobank. The upper panel shows deletion and duplication carrier frequencies at the lead probe for each locus, derived from the mirror dosage matrix across all 94,793 QC-passing participants. The lower panel shows CNV-wide significant cytoband–trait associations, coloured according to CNV model class. Horizontal dashed lines indicate the CNV-wide significance thresholds (P ≤ 8.40 × 10⁻ for deletions, P ≤ 1.33 × 10⁻ for duplications, and P ≤ 9.24 × 10⁻ for mirror models). Text labels indicate the direction of effect on the inverse rank-normal transformed trait. Traits with no CNV-wide significant associations (heart rate, BMI at age 25, and diastolic blood pressure) are not shown. Cytobands are ordered by chromosome. Where a single cytoband harbours two distinct loci, these are distinguished by the suffixes a and b according to genomic position: 1q21.1a (142.9 Mb), 1q21.1b (146.7 Mb), 7p21.1a (17.5 Mb), and 7p21.1b (20.0 Mb) (lead probe positions; hg19 reference genome build).

**Table 1.** CNV loci associated with anthropometric and cardiometabolic traits in the China Kadoorie Biobank.

| Locus | Phenotype | Start<br>(Mb) | End<br>(Mb) | Del<br>(n) | Dup<br>(n) | Model | $\beta$ | P |
| --- | --- | --- | --- | --- | --- | --- | --- | --- |
| 1q12 | Resting heart rate | 142.5 | 142.5 | 254 | - | Del | -0.30 | $2.27 \times 10^{-6}$ |
| 1q21.1 | RPG | 142.7 | 143.0 | 461 | - | Del | -0.22 | $3.67 \times 10^{-6}$ |
| 1q21.1 | Standing height | 146.6 | 147.4 | 28 | 21 | Del, Mirror | +0.46 | $1.92 \times 10^{-6}$ |
| 2q22.1 | RPG | 137.6 | 137.6 | 2 | 22 | Mirror | -0.91 | $8.02 \times 10^{-6}$ |
| 7p21.1 | Resting heart rate | 17.5 | 17.5 | 24 | - | Del | -0.94 | $3.72 \times 10^{-6}$ |
| 7p21.1 | DBP | 20.0 | 20.0 | 24 | 13 | Mirror | +0.75 | $5.25 \times 10^{-6}$ |
| 8p23.1 | RPG | 12.4 | 12.5 | 361 | - | Del | -0.29 | $5.40 \times 10^{-8}$ |
| 8p22 | Sitting height | 13.8 | 13.8 | 33 | 16 | Mirror | +0.45 | $4.39 \times 10^{-6}$ |
| 12p12.1 | Fat percentage | 24.9 | 25.1 | - | 23 | Dup | -0.74 | $8.11 \times 10^{-6}$ |
| 14q11.2 | RPG | 22.8 | 23.0 | 111 | 19 | Del, Mirror | +0.43 | $8.41 \times 10^{-7}$ |
| 15q13.1 | Standing height | 28.2 | 28.2 | 4 | 13 | Mirror | -0.73 | $8.18 \times 10^{-6}$ |
| 16p13.2 | Standing height | 10.4 | 10.4 | 3 | 13 | Mirror | -0.75 | $7.49 \times 10^{-6}$ |
| 16p11.2<sup>a</sup> | BMI | 28.8 | 29.0 | 8 | 35 | Dup, Mirror | -0.84 | $1.77 \times 10^{-8}$ |
| | Fat percentage | 28.8 | 29.0 | - | 35 | Dup | -0.63 | $2.52 \times 10^{-6}$ |
| | Hip circumference | 28.8 | 29.0 | 8 | 38 | Dup, Mirror | -0.69 | $2.34 \times 10^{-7}$ |
| | Waist circumference | 28.8 | 29.0 | 8 | 35 | Dup, Mirror | -0.74 | $5.02 \times 10^{-7}$ |
| | Weight | 28.8 | 29.0 | 8 | 38 | Dup, Mirror | -0.67 | $1.84 \times 10^{-7}$ |
| 16p11.2<sup>b</sup> | Fat percentage | 29.6 | 30.2 | 12 | 29 | Dup, Mirror | -0.72 | $5.34 \times 10^{-9}$ |
| 17q12 | Fat percentage | 34.4 | 34.4 | - | 40 | Dup | -0.56 | $7.36 \times 10^{-6}$ |

All positions are hg19 (Mb). $\beta$ , effect size in SD units of the inverse rank-normal transformed phenotype; for mirror models, $\beta$ represents the per-copy effect (duplication +1, deletion -1). "-" indicates a CNV type not applicable under the reported model. Where both a single-type and mirror model reached CNV-wide significance, mirror model statistics are reported; the concordant single-type model is indicated in the Model column. Full results for all models are in Tables S3 and S4. RPG, random plasma glucose; DBP, diastolic blood pressure; BMI, body mass index.
<sup>a</sup> 16p11.2 BP2–BP3 segment (~28.8–29.0 Mb).
<sup>b</sup> 16p11.2 BP4–BP5 segment (~29.6–30.2 Mb)

#### Adiposity trait associations at 16p11.2 loci

A prominent multi-trait locus mapped to 16p11.2 with a lead probe intragenic to *SH2B1* (MIM: 608937; lead position ∼28.865 Mb) and a CNVR spanning chr16:28.78-29.01 Mb (**Figure 4A**). *SH2B1* encodes an adaptor protein in leptin, insulin, and growth hormone signalling; loss-of-function mutations have been associated with severe early-onset obesity and hyperphagia^50^. This interval overlaps the recurrent 16p11.2 BP2-BP3 CNV region (deletion syndrome MIM: 613444), previously implicated in early-onset obesity and neurodevelopmental phenotypes^51,52^. In our cohort, signals at this locus were detected under both duplication-only and mirror encodings across multiple adiposity traits. Duplications (35 carriers; 0.037%) were associated with lower BMI (β = −0.886, SE = 0.165, *P* = 7.97×10⁻), lower weight (β = −0.836, SE = 0.147, *P* = 1.39×10⁻), smaller WC (β = −0.766, SE = 0.162, p = 2.29×10⁻), smaller HC (β = −0.774, SE = 0.146, *P* = 1.24×10⁻), and lower body fat percentage (β = −0.630, SE = 0.134, *P* = 2.52×10⁻) (**Table 1; Table ST4**). Mirror-model analyses at the same locus (43 carriers; 8 deletions and 35 duplications; 0.045%) also reached experiment-wide significance for BMI (β = −0.839, SE = 0.149, *P* = 1.77×10⁻), HC (β = −0.688, SE = 0.133, *P* = 2.34×10⁻), WC (β = −0.735, SE = 0.146, *P* = 5.02×10⁻), and weight (β = −0.670, SE = 0.129, *P* = 1.84×10⁻) (**Table 1; Table ST5**). Unadjusted mean (median) BMI was 25.7 (25.4) kg/m² in deletion carriers, 23.7 (23.4) kg/m² in copy-neutral individuals, and 20.8 (20.5) kg/m² in duplication carriers (**Figure 4B**), consistent with a bidirectional dosage effect. Associations between BP2-BP3 CNVs and adiposity traits have been previously reported in both clinical^51^ and large-scale CNV-GWAS in European cohorts^13,16–18^. This CNVR contained dense GWAS Catalog support for adiposity traits, consistent with shared dosage-sensitive biology at this locus.

**Figure 4.**
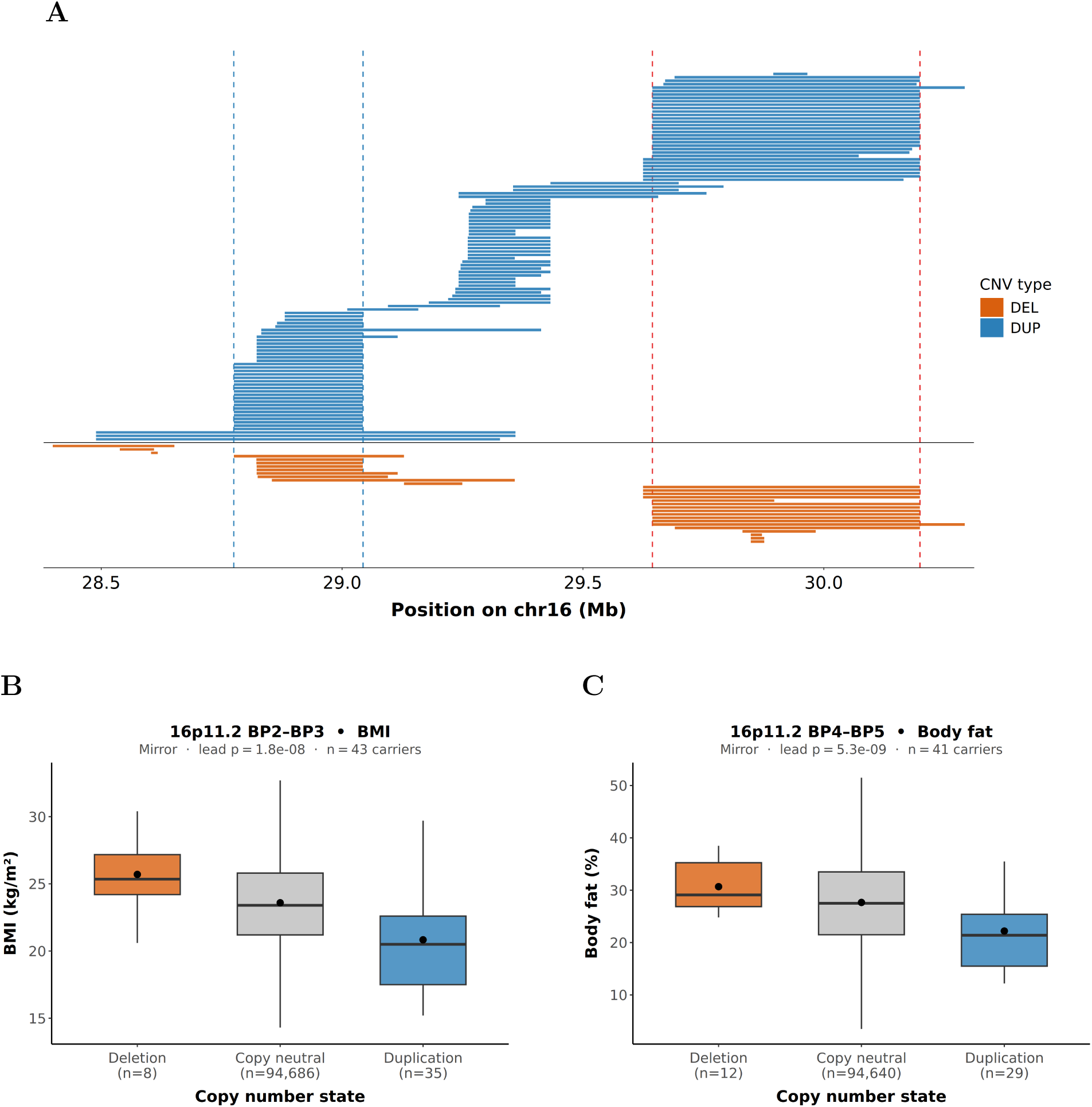
16p11.2 CNV associations with adiposity traits. (A) CNV distribution across 16p11.2 hg19, showing proximal and distal recurrent CNV clusters. Blue vertical lines indicate CNVR boundaries for the proximal region in CKB, and red vertical lines indicate CNVR boundaries for the distal region. (B) Mirror-model boxplot showing BMI association at the proximal locus. Mean (median) BMI was 25.7 (25.4) in deletion carriers, 23.7 (23.4) in copy-neutral individuals, and 20.8 (20.5) in duplication carriers. (C) Mirror-model boxplot showing body fat percentage association at the distal locus. Mean (median) body fat percentage was 30.7% (29.1%) in deletion carriers, 27.7% (27.5%) in copy-neutral individuals, and 22.2% (21.4%) in duplication carriers.

A second, distinct interval within 16p11.2, separable from the *SH2B1* locus by both coordinates and gene content, was also associated with body fat percentage (**Figure 4C**). This locus is centred on a lead probe at chr16:30,174,024 and spans a CNVR of chr16:29.64-30.20 Mb, overlapping the recurrent 16p11.2 BP4-BP5 CNV region (deletion MIM: 611913; duplication MIM: 614671). The association was detected under both duplication-only (β = −0.765, SE = 0.147, *P* = 1.94×10⁻; 29 duplication carriers; 0.031%) and mirror (β = −0.721, SE = 0.124, *P* = 5.34×10⁻; 12 deletions and 29 duplications; 41 total carriers; 0.043%) models (**Table 1; Table ST4; Table ST5**). Unadjusted mean (median) body fat percentage was 30.7% (29.1%) in deletion carriers, 27.7% (27.5%) in copy-neutral individuals, and 22.2% (21.4%) in duplication carriers (**Figure 4C**), with opposing directions of effect consistent with a bidirectional dosage architecture, though deletion carrier numbers were small (n = 12). The opposing directions of effect in deletion and duplication carriers are consistent with the bidirectional dosage architecture at this locus first characterised in clinical cohorts^51,53,54^, and subsequently replicated in large-scale CNV-GWAS in European populations^13,16–18^; our findings extend this association to a Chinese population for the first time. This CNVR also showed dense GWAS Catalog support for adiposity traits.

#### Body fat percentage associations at 12p12.1 and 17q12

Two additional duplication-associated reductions in body fat percentage were identified at loci not previously reported in CNV association studies. One locus mapped to a 300 kb region on 12p12.1 (lead probe chr12:25,047,720; *P* = 8.11×10⁻; β = −0.737, SE = 0.165; 23 carriers, 0.024%, **Figure 5A**; **Table 1**; **Table ST4**), with the lead probe falling within *BCAT1* (MIM: 113520). The tagged CNVR (chr12:24.92-25.23 Mb) spans *BCAT1*, *AC026310.1*, *C12orf77*, and *LRMP*, with carrier CNVs additionally overlapping *CASC1* at the distal edge. *BCAT1* encodes a key enzyme in branched-chain amino acid (BCAA) metabolism^55^, and circulating BCAAs are strongly associated with adiposity, insulin resistance, and incident type 2 diabetes^56,57^. The observed direction of effect, lower body fat percentage in duplication carriers, is therefore consistent with altered dosage of a metabolically relevant gene, although direct functional evidence at the CNV level is lacking. Unadjusted mean (median) body fat percentage was 19.9% (17.0%) in duplication carriers compared with 27.7% (27.5%) in copy-neutral individuals (**Figure 5A**). No trait-matched GWAS Catalog SNP associations fell within the tagged CNVR.

**Figure 5.**
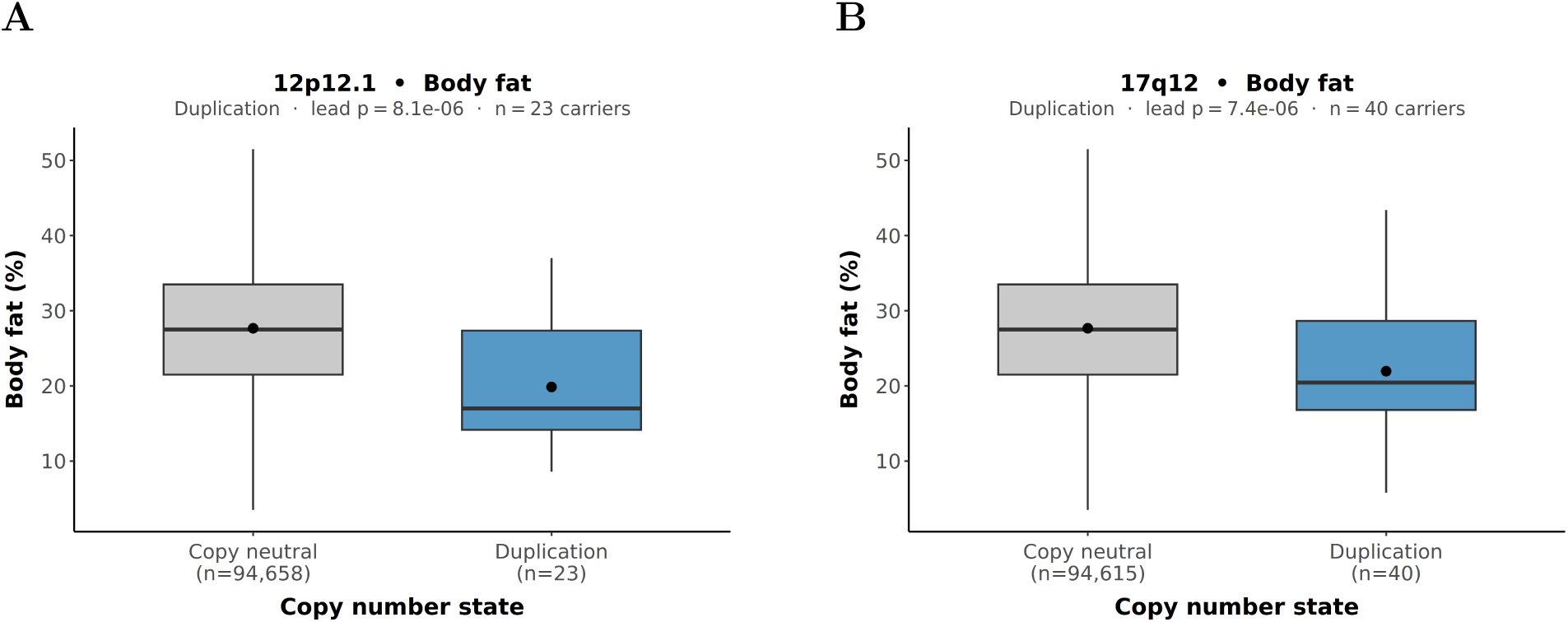
Duplications at 12p12.1 and 17q12 are associated with lower body fat percentage. (A) Duplication-model boxplot showing body fat percentage association at 12p12.1. Mean (median) body fat was 19.9% (17.0%) in duplication carriers (n=23) and 27.7% (27.5%) in copy-neutral individuals. (B) Duplication-model boxplot showing body fat percentage association at 17q12. Mean (median) body fat was 22.0% (20.5%) in duplication carriers (n=40) and 27.7% (27.5%) in copy-neutral individuals.

A further locus mapped to a small (3.4 kb) region on 17q12 (lead probe chr17:34,422,135; *P* = 7.36×10⁻; β = −0.561, SE = 0.125; 40 carriers, 0.042%, **Figure 5B**; **Table 1; Table ST4**), approximately 400 kb proximal to the well-characterised 17q12/*HNF1B* recurrent deletion interval (deletion MIM: 614527; duplication MIM: 614526). The canonical interval, which our CNVR does not overlap, harbours *HNF1B* and has been associated with renal anomalies, maturity-onset diabetes of the young type 5 (MODY5), and neurodevelopmental phenotypes in deletion carriers, and variable neurodevelopmental features in duplication carriers^58,59^. The CNVR (chr17:34.42-34.43 Mb) spans no annotated protein-coding genes, although the lead probe lies proximal to a structurally complex region containing *TBC1D3* paralogues and members of the *CCL3/CCL4* chemokine cluster^59^. Unadjusted mean (median) body fat percentage was 22.0% (20.5%) in duplication carriers compared with 27.7% (27.5%) in copy-neutral individuals (**Figure 5B**). No trait-matched GWAS Catalog associations were identified within the CNVR. A mirror-model signal was also identified at this locus but had zero deletion carriers at the lead probe and is therefore not reported separately.

#### Diastolic blood pressure associations at 7p21.1

DBP was associated at 7p21.1 under the mirror model (CNVR chr7:19,960–20,011 kb; β = +0.750, SE = 0.165, *P* = 5.25×10⁻; 37 carriers: 24 deletions, 13 duplications; **Table 1**; **Table ST5**). Deletion CNVs were short (median 38 kb) whereas duplication CNVs were substantially longer (median 654 kb), with the two CNV types showing markedly different genomic extents at this locus; independent effects specific to each type cannot therefore be excluded. Unadjusted mean DBP was 70.0 mmHg in deletion carriers, 78.6 mmHg in copy-neutral individuals, and 84.9 mmHg in duplication carriers (**Figure 6**). The lead probe falls in an intergenic region flanked by *TWISTNB* and *MACC1*; no trait-matched GWAS Catalog associations were identified, and a well-established BP-associated cluster within *HDAC9*^60,61^ lies ∼940 kb proximal, within range only of the single longest duplication carrier. Deletion carriers at a second, more proximal 7p21.1 locus (CNVR chr7:17.5 Mb; β = −0.943, SE = 0.204, *P* = 3.72×10⁻; 24 carriers) were also associated with lower resting heart rate (**Table 1**; **Table ST4**); this locus is distinct from the DBP locus (∼2.5 Mb distal) and has no annotated biological correlate or trait-matched GWAS Catalog support.

**Figure 6.**
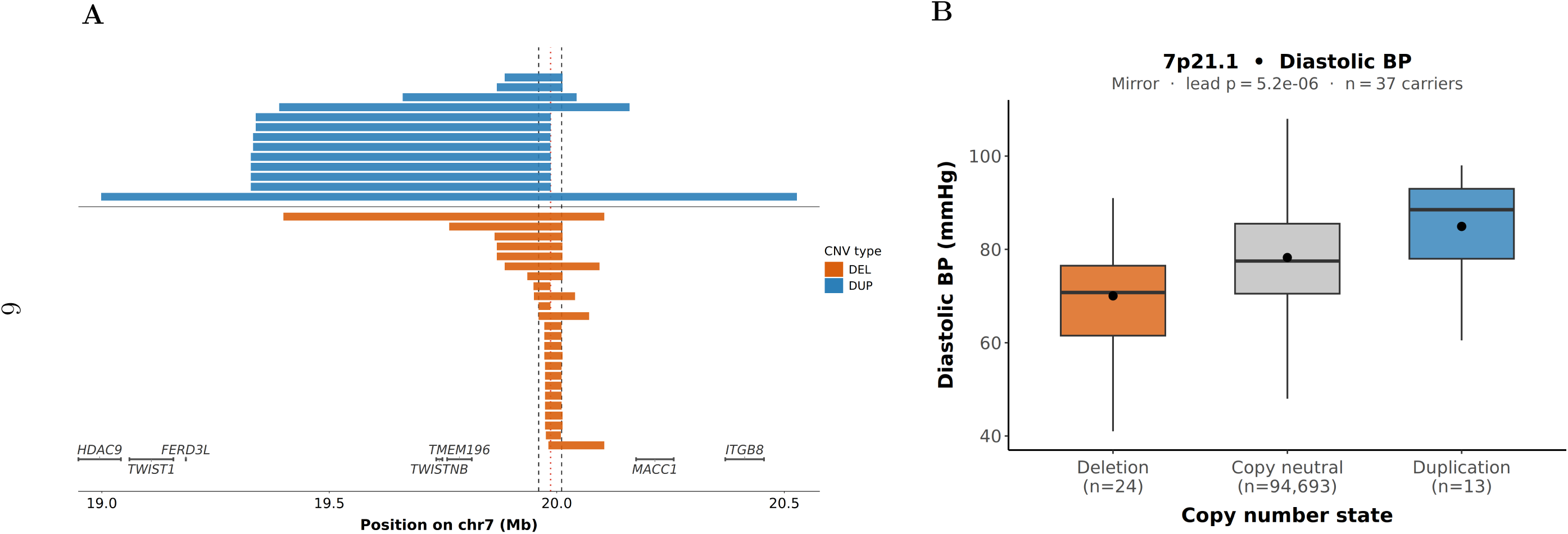
7p21.1 CNV association with DBP. (A) CNV distribution across 7p21.1 (hg19), showing deletion and duplication carriers overlapping the CNVR interval. Black dashed lines indicate CNVR boundaries and the red dotted line indicates the lead probe position. (B) Mirror-model boxplot showing DBP association at 7p21.1. Mean (median) DBP was 70.0 (70.8) mmHg in deletion carriers (n = 24), 78.6 (77.5) mmHg in copy-neutral individuals, and 84.9 (88.5) mmHg in duplication carriers (n = 13).

#### Random plasma glucose associations

The strongest RPG signal mapped to 8p23.1 (CNVR chr8:12.37–12.53 Mb; deletion model; β = −0.285, SE = 0.052, *P* = 5.40×10⁻; 361 carriers, 0.386%), within a structurally complex region harbouring β-defensin genes and predisposed to recurrent rearrangement by REPD/REPP segmental duplication architecture^46^. This locus has previously been implicated in type 2 diabetes through linkage analysis^62^, and the association remained CNV-wide significant after adjustment for time since last meal (*P* = 1.80×10⁻; **Table 1**; **Table ST4**). A deletion signal at 1q21.1 (CNVR chr1:142.54–144.02 Mb; β = −0.215, *P* = 3.67×10⁻; 461 carriers) was attenuated after meal-time adjustment (*P* = 1.96×10⁻); carrier-level analysis confirmed near-complete overlap (252/254 carriers) with the pericentromeric 1q12 deletion driving the resting heart rate association, consistent with the same underlying structural events contributing to both traits.

A mirror-model signal at 14q11.2 (CNVR chr14:22.75–22.95 Mb; β = +0.430, SE = 0.087, *P* = 8.41×10⁻; 130 carriers: 111 deletions, 19 duplications) showed a bidirectional dosage pattern, with deletion carriers showing lower and duplication carriers higher RPG relative to copy-neutral individuals; this association survived fasting time adjustment (**Table 1; Tables ST3–ST4**). However, this CNVR overlaps the T-cell receptor delta/alpha locus, which undergoes somatic V(D)J recombination in T lymphocytes; as genotyping was performed on blood-derived DNA, apparent CNV calls here may partly reflect somatic rearrangements rather than germline structural variation^63^, and this association should be interpreted with caution. A further mirror-model signal at 2q22.1 (CNVR chr2:137.24–137.60 Mb; β = −0.906, *P* = 8.02×10⁻; 24 carriers: 2 deletions, 22 duplications) implicates THSD7B but rests on only 2 deletion carriers; duplication-only analyses across the region showed consistent negative associations (effect sizes −0.64 to −0.90), suggesting a genuine duplication-driven effect that narrowly missed the duplication-model threshold (**Table 1; Table ST4**).

#### Resting heart rate association at 1q12

Resting heart rate was associated with deletions at 1q12 (CNVR chr1:142.54–143.42 Mb; β = −0.297, SE = 0.063, *P* = 2.27×10⁻; 254 deletion carriers, 0.269%; **Table 1; Table ST4**). As noted above, 252 of these 254 carriers also carried the 1q21.1 RPG-associated deletion, consistent with the same pericentromeric events driving both associations. No trait-matched GWAS Catalog associations were identified.

#### Standing and sitting height associations

Standing height was associated at 1q21.1 under both deletion-only (CNVR chr1:146.50–147.55 Mb; β = −0.513, SE = 0.105, *P* = 1.04×10⁻; 41 deletion carriers, 0.043%) and mirror (β = +0.458, SE = 0.096, *P* = 1.92×10⁻; 49 carriers: 28 deletions, 21 duplications) models (**Table 1; Tables ST3–ST4).** This CNVR corresponds to the recurrent 1q21.1 deletion (MIM: 612474) and duplication (MIM: 612475) syndromes, previously associated with altered growth and skeletal phenotypes^64^, and is distinct from the more proximal 1q12 locus implicated for RPG and heart rate. The CNVR overlaps GWAS Catalog SNP associations for height^65^, and replicates CNV-height signals reported under mirror models in both a large European CNV meta-analysis^16^ and a UK Biobank CNV-GWAS^17^. Two further mirror-model associations for standing height were identified: at 15q13.1 (CNVR chr15:26.85–28.33 Mb; lead probe intragenic to *OCA2* [MIM: 611409]; β = −0.728, *P* = 8.18×10⁻; 17 carriers: 4 deletions, 13 duplications; two GWAS Catalog height RSIDs within CNVR) and at 16p13.2 (lead probe intragenic to *ATF7IP2*; β = −0.753, *P* = 7.49×10⁻; 16 carriers: 3 deletions, 13 duplications). The 16p13.2 signal is unlikely to reflect a genuine dosage effect: it is attributable to the extreme stature of 3 deletion carriers (mean 173.5 cm versus 158.6 cm in copy-neutral individuals), with no corresponding signal in duplication-only analyses; it is reported for completeness (**Table 1; Table ST5**).

Sitting height was associated at 8p22 under the mirror model (CNVR chr8:13.76–13.86 Mb; β = +0.455, SE = 0.099, *P* = 4.39×10⁻; 49 carriers: 33 deletions, 16 duplications; no GWAS Catalog support; **Table 1**; **Table ST5**).

#### Sensitivity analyses among unrelated individuals

Sensitivity analyses restricted to individuals with no first-to third-degree relatives in the dataset (n = 71,477–72,183 depending on trait) showed consistent results across all 26 model-specific associations meeting CNV-wide significance thresholds. Effect estimates were directionally concordant between the full and unrelated analyses for all 26 signals, with closely concordant effect sizes (**Figure S6**). For all 26 associations, the lead probe ranked within the top 5% of probe-level *p*-values for the corresponding phenotype and model in the unrelated-only analysis, and within the top 1% for 24 of 26. Eight signals remained CNV-wide significant under unrelated-only thresholds, while the remaining signals showed consistent prioritisation but did not exceed model-specific thresholds, consistent with reduced power in the smaller unrelated subset.

## Discussion

Large-scale CNV association studies remain substantially under-represented in non-European populations, limiting understanding of the extent to which dosage-sensitive loci generalise across ancestries or reflect population-specific CNV architectures^66^. This study of 94,730 Chinese adults broadens the ancestral diversity represented in large-scale CNV research. Genome-wide CNV association analyses across 13 quantitative traits revealed 19 locus-phenotype associations. These include signals not previously reported in CNV studies, for example, CNVRs at 12p12.1 and 17q12 for body fat percentage, 7p21.1 for DBP, and 8p23.1 for RPG. The candidate novel locus-phenotype associations identified in this study expand the known contribution of structural variation to anthropometric and cardiometabolic trait biology beyond that previously detected in predominantly European ancestry CNV-GWAS.

Findings from the present study included several well-established dosage-sensitive loci previously observed largely in European ancestry populations. The most prominent replicated finding was a cluster of associations at 16p11.2 spanning 5 adiposity traits across both the BP2-BP3 interval overlapping SH2B1 and the BP4-BP5 interval. Both loci have been robustly associated with adiposity in previous studies^9,16,17,53,54^, and the bidirectional dosage pattern is consistent with previously reported mirror effects^17,53^. To our knowledge, these represent the first CNV-wide significant associations between 16p11.2 CNVs and adiposity traits in an East Asian population. By contrast, a prior candidate study in a Han Chinese population returned a null finding at this locus. However, that likely reflects limited statistical power given the previous study’s much smaller sample size^67^.

The associations observed with adiposity in the present study at these established loci suggest dosage sensitivity at these loci generalises across populations with markedly different adiposity profiles. For example, the estimated effect of the BP2-BP3 duplication on BMI in CKB (-3.10 kg/m²), in which the mean BMI was 23.7 kg/m², was comparable in magnitude to deletion effects reported in European cohorts (+3.07 kg/m² in Macé et al., 2017^16^; +4.25 kg/m² in Auwerx et al., 2022^17^), in which average BMI levels were notably higher (for example, 27.4 kg/m^2^ in the Auwerx et al study population^17^). A standing height association at 1q21.1 similarly replicated previously reported CNV-height associations^17,64^, with concordant SNP-GWAS support reinforcing the signal at this locus. Moreover, annotation of CNVRs with published SNP-GWAS associations from the NHGRI-EBI GWAS Catalog^42^ identified dense same-trait SNP signals at some established loci (eg, 16p11.2 and 1q21.1 for standing height), consistent with shared dosage-sensitive biology. Although, several established CNV-trait associations, including signals at 22q11.21 for BMI^10,68^, were not recovered in this study, this likely reflects reduced power in population-based cohorts, where carriers of highly penetrant CNVs are underrepresented, together with known differences in CNV frequency across ancestry groups^16,30^.

Two of the candidate novel loci identified in the present study were associated with body fat percentage. Notably, these were not found to be associated with other measures of adiposity in our study, including BMI. Well-established differences in adiposity patterns exist between East Asian and European populations. East Asian ancestry populations show a propensity to higher levels of body fat at equivalent BMI^69,70^ and greater central adiposity^71^ compared with individuals of European ancestry despite a typically leaner phenotype^69,71^. These differences may partly explain why the candidate loci identified in the present study were associated with body fat percentage rather than BMI. More broadly, population-specific differences in adiposity biology may contribute to the identification of loci not previously reported in predominantly European ancestry CNV studies. However, the absence of prior associations at these loci may also reflect differences in CNV frequency across ancestries or limited statistical power in previous studies. Additional candidate novel loci were identified for RPG, a phenotype which also exhibits differences between populations. For example, in East Asian populations, post-load glucose dysregulation is an earlier and more prominent feature of dysglycaemia than in Europeans, where impaired fasting glucose is comparatively more predominant^72^. RPG may therefore represent a particularly sensitive phenotype for detecting glycaemic loci in the CKB population. Consistent with this, four RPG-associated CNVRs were identified, including a deletion signal at 8p23.1 that remained significant after adjustment for time since last meal and exceeded the experiment-wide significance threshold, a highly conservative benchmark given the substantial inter-probe correlation structure across CNV regions. Moreover, this signal maps to a structurally complex region previously implicated in type 2 diabetes linkage studies^62^, lending biological plausibility to these findings. Replication in independent East Asian cohorts and functional follow-up will be needed to confirm and further characterise these associations.

Annotation of CNVRs with published SNP-GWAS associations from the NHGRI-EBI GWAS Catalog^42^ identified same-trait SNP signals at 16p11.2 for adiposity traits, and both 1q21.1 and 15q13.1 for standing height. At all other reported loci, however, SNP support was limited or absent within the corresponding r²-defined intervals. This should be interpreted cautiously: CNVs and flanking SNPs are known to be in significantly lower linkage disequilibrium than SNP pairs, particularly for duplications^73,74^, such that genuine CNV effects may not be adequately captured by nearby array SNPs even within the same locus. SNP annotation is therefore presented here primarily as contextualisation rather than evidence for shared or independent mechanisms. More broadly, the incomplete overlap between several CNVRs and established SNP-GWAS loci supports growing evidence that structural variation contributes components of complex trait architecture not readily detectable through SNP-based association studies alone^66^.

The present study has several notable strengths. These analyses in an East Asian population enabled evaluation of both established dosage-sensitive loci and candidate novel associations in an ancestry group substantially underrepresented in previous CNV-GWAS. The combination of probe-level association testing, array-stratified quality control, and sensitivity analyses in unrelated participants further supports the robustness of the reported signals. However, this study also has some limitations. Probe-level association testing, as applied here and in previous large-scale CNV-GWAS^16,17^, allows partially overlapping CNV events with heterogeneous breakpoints to contribute jointly to association signals without requiring exact breakpoint concordance. The r²-defined CNVRs reported here should therefore be interpreted as association-defined intervals rather than precise representations of any single structural event. CNVs are rare: fewer than 0.2% of probes exceeded a carrier frequency of 1%, and statistical power was consequently concentrated at recurrent hotspots. The minimum carrier threshold of ≥15 serves a role analogous to a minor allele frequency filter in SNP-GWAS, reducing instability in regression estimates at the cost of excluding very rare events. Events occurring in regions of low probe density may be missed entirely, and non-differential misclassification of CNV status would be expected to attenuate effect estimates toward the null^75^, suggesting that the associations identified here likely represent only a subset of the true CNV-trait relationships detectable at biobank scale. These limitations will be reduced as whole-genome sequencing becomes available for CKB participants, enabling variant-level and gene-level burden analyses with improved sensitivity and breakpoint resolution^15,18,76^.

Several study-specific limitations should also be noted. The main analyses included related individuals. However, sensitivity analyses restricted to unrelated participants confirmed directional concordance across all 26 model-specific signals. Array-stratified analyses additionally demonstrated consistency of effect estimates across the two genotyping platforms, arguing against systematic platform-specific artefacts. Carrier numbers nevertheless remained small at several loci, and these associations should therefore be regarded as statistically supported candidate signals pending replication. No independent East Asian cohort with comparable CNV data was available for external validation. Finally, as a single-ancestry study, findings may not generalise to other East Asian subgroups or to non-East-Asian populations. Comparisons with other ancestries will be important to determine the extent to which identified CNV-trait associations reflect population-specific or broadly shared effects.

In conclusion, this study provides the largest genome-wide CNV association analysis of quantitative traits in an East Asian population to date, identifying 19 locus-phenotype associations across anthropometric and cardiometabolic traits in 94,730 Chinese adults. Alongside replication of well-established dosage-sensitive loci, candidate novel signals across a range of cardiometabolic loci highlight that the CNV contribution to complex trait variation in Chinese adults extends beyond what has been characterised in predominantly European cohorts. Together, these findings support emerging efforts to extend genome-wide association analyses beyond SNP-centric models toward more comprehensive incorporation of structural variation into the genetic architecture of complex traits^76^. Improved characterisation of dosage-sensitive regions across diverse populations may ultimately contribute to more complete models of disease susceptibility, more accurate causal gene mapping, and identification of biologically relevant pathways for future therapeutic investigation.

## Data Availability

The China Kadoorie Biobank (CKB) is a global resource for the investigation of lifestyle, environmental, biochemical and genetic factors as determinants of common diseases. Non-genetic data (e.g., baseline, resurveys, biomarkers, and disease endpoints) are released periodically to bona fide researchers. Details of the CKB Data Sharing Policy, data release schedules and formal application procedures are available at www.ckbiobank.org/data-access. Access to individual participant genetic data is currently constrained by the Administrative Regulations on Human Genetic Resources of the People's Republic of China and is usually facilitated through collaboration with CKB researchers; researchers interested in accessing the underlying genetic data should contact, where a research proposal will be requested. Code used for the analyses in this study can be made available by contacting the corresponding authors.

## Acknowledgements

The most important acknowledgement is to the participants in the study and the members of the survey teams in each of the 10 regional centres, as well as to the project development and management teams based at Beijing, Oxford and the 10 regional centres. We thank Hongcheng Zhou, Haoxiang Lin, Jieqin Liang, and their colleagues at BGI (Shenzhen, China) for DNA extraction, genotyping array design, and genotyping. The CKB baseline survey and the first re-survey were supported by the Kadoorie Charitable Foundation in Hong Kong. The long-term follow-up has been supported by Wellcome grants to Oxford University (212946/Z/18/Z, 202922/Z/16/Z, 104085/Z/14/Z, 088158/Z/09/Z) and grants from the National Natural Science Foundation of China (82192901, 82192904, 82192900, 82388102) and the Noncommunicable Chronic Diseases-National Science and Technology Major Project (2023ZD0510101, 2023ZD0510100).The UK Medical Research Council (MC_UU_00017/1, MC_UU_12026/2, MC_U137686851), Cancer Research UK (C16077/A29186; C500/A16896) and the British Heart Foundation (CH/1996001/9454), provide core funding to the Clinical Trial Service Unit and Epidemiological Studies Unit at Oxford University for the project. DNA extraction and genotyping was funded by GlaxoSmithKline and the UK Medical Research Council (MC-PC-13049, MC-PC-14135). IH was supported by the Oxford EPSRC Centre for Doctoral Training in Health Data Science (EP/S02428X/1). FB was supported by Health Data Research UK which is funded by UK Research and Innovation, the Medical Research Council, the British Heart Foundation, Cancer Research UK, the National Institute for Health and Care Research, the Economic and Social Research Council, the Engineering and Physical Sciences Research Council, Health and Care Research Wales, Health and Social Care Research and Development Division (Public Health Agency, Northern Ireland), Chief Scientist Office of the Scottish Government Health and Social Care Directorates. This research was funded in whole, or in part, by the Wellcome Trust (212946/Z/18/Z, 202922/Z/16/Z, 104085/Z/14/Z, 088158/Z/09/Z). For the purpose of Open Access, the author has applied a CC-BY public copyright licence to any Author Accepted Manuscript version arising from this submission.

## Author contributions

I.H. and R.G.W. analysed the data. I.H. drafted the manuscript. Z.C., R.G.W., F.B. and D.B. contributed to the conception of this paper, interpretation of the results, and revision of the manuscript. S.M., K.L., I.M., C.Y., J.L., D.S., and Z.C. provided administrative, technical, and research support, including oversight of data collection and cohort infrastructure. D.A. contributed to data processing and quality control. All authors critically reviewed the manuscript and approved the final submission.

