## Supplementary Figures for "Copy number variant association analysis in 94,730 Chinese adults reveals loci influencing anthropometric and cardiometabolic traits"

### Supplementary Materials


Members of the China Kadoorie Biobank Collaborative Group 2

Supplementary Note 1: Array Design 4

Figure S1 Study flowchart 5

Figure S2 Distribution of CNV calling quality control metrics across CKB genotyped population 6

Figure S3 Array-version-stratified and pooled effect estimates at each lead probe–phenotype association 7

Figure S4 Lead probe carrier counts at each reported association, stratified by array version 8

Figure S5 CNV×array version interaction significance versus CKB1 carrier count at the lead probe 9

Figure S6 Unadjusted CNV carrier group distributions for BMI at 16p11.2 BP2–BP3 10

Figure S7 Unadjusted CNV carrier group distributions for diastolic blood pressure at 7p21.1 11

Figure S8 Unadjusted CNV carrier group distributions for body fat percentage at 12p12.1 12

Figure S9 Unadjusted CNV carrier group distributions for body fat percentage at 16p11.2 BP4–BP5 13

Figure S10 Unadjusted CNV carrier group distributions for body fat percentage at 16p11.2 BP2–BP3 14

Figure S11 Unadjusted CNV carrier group distributions for body fat percentage at 17q12 15

Figure S12 Unadjusted CNV carrier group distributions for resting heart rate at 1q12 16

Figure S13 Unadjusted CNV carrier group distributions for resting heart rate at 7p21.1 17

Figure S14 Unadjusted CNV carrier group distributions for hip circumference at 16p11.2 BP2–BP3 18

Figure S15 Unadjusted CNV carrier group distributions for random glucose at 1q21.1 19

Figure S16 Unadjusted CNV carrier group distributions for random glucose at 2q22.1 20

Figure S17 Unadjusted CNV carrier group distributions for random glucose at 8p23.1 21

Figure S18 Unadjusted CNV carrier group distributions for random glucose at 14q11.2 22

Figure S19 Unadjusted CNV carrier group distributions for sitting height at 8p22 23

Figure S20 Unadjusted CNV carrier group distributions for standing height at 1q21.1 24
Figure S21 Unadjusted CNV carrier group distributions for standing height at 15q13.1 25
Figure S22 Unadjusted CNV carrier group distributions for standing height at 16p13.2 26
Figure S23 Unadjusted CNV carrier group distributions for waist circumference at 16p11.2 BP2–BP3 27
Figure S24 Unadjusted CNV carrier group distributions for weight at 16p11.2 BP2–BP3 28
Figure S25 Concordance of effect estimates between the full sample and an unrelated subsample 29

#### Members of the China Kadoorie Biobank Collaborative Group

**International Steering Committee:** Junshi Chen, Zhengming Chen (PI), Robert Clarke, Rory Collins, Liming Li (PI), Jun Lv, Richard Peto, Robin Walters.

**International Co-ordinating Centre, Oxford:** Daniel Avery, Maxim Barnard, Derrick Bennett, Ruth Boxall, Ka Hung Chan, Yiping Chen, Zhengming Chen, Charlotte Clarke, Jonathan Clarke, Robert Clarke, Huaidong Du, Ahmed Edris Mohamed, Hannah Fry, Simon Gilbert, Pek Kei Im, Andri Iona, Maria Kakkoura, Christiana Kartsonaki, Hubert Lam, Kuang Lin, James Liu, Mohsen Mazidi, Iona Millwood, Sam Morris, Qunhua Nie, Alfred Pozarickij, Maryam Rahmati, Paul Ryder, Dan Schmidt, Becky Stevens, Iain Turnbull, Robin Walters, Baihan Wang, Lin Wang, Neil Wright, Ling Yang, Xiaoming Yang, Pang Yao.

**National Co-ordinating Centre, Beijing:** Xiao Han, Can Hou, Qingmei Xia, Chao Liu, Jun Lv, Pei Pei, Dianjanyi Sun, Canqing Yu, Lang Pan.

**10 Regional Co-ordinating Centres:**

**Qingdao CDC:** Zengchang Pang, Ruqin Gao, Shanpeng Li, Haiping Duan, Shaojie Wang, Yongmei Liu, Ranran Du, Yajing Zang, Liang Cheng, Xiaocao Tian, Hua Zhang, Yaoming Zhai, Feng Ning, Xiaohui Sun, Feifei Li. **Licang CDC:** Silu Lv, Junzheng Wang, Wei Hou. **Heilongjiang Provincial CDC:** Wei Sun, Shichun Yan, Xiaoming Cui. **Nangang CDC:** Chi Wang, Zhenyuan Wu, Yanjie Li, Quan Kang. **Hainan Provincial CDC:** Huiming Luo, Tingting Ou. **Meilan CDC:** Xiangyang Zheng, Zhendong Guo, Shukuan Wu, Yilei Li, Huimei Li. **Jiangsu Provincial CDC:** Ming Wu, Yonglin Zhou, Jinyi Zhou, Ran Tao, Jie Yang, Jian Su. **Suzhou CDC:** Fang Liu, Jun Zhang, Yihe Hu, Yan Lu, Liangcai Ma, Aiyu Tang, Shuo Zhang, Jianrong Jin, Jingchao Liu. **Guangxi Provincial CDC:** Mei Lin, Zhenzhen Lu. **Liuzhou CDC:** Lifang Zhou, Changping Xie, Jian Lan, Tingping Zhu, Yun Liu, Liuping Wei, Liyuan Zhou, Ningyu Chen, Yulu Qin, Sisi Wang. **Sichuan Provincial CDC:** Xianping Wu, Ningmei Zhang, Xiaofang Chen, Xiaoyu Chang. **Pengzhou CDC:** Mingqiang Yuan, Xia Wu, Xiaofang Chen, Wei Jiang, Jiaqiu Liu, Qiang Sun. **Gansu Provincial CDC:** Faqing Chen, Xiaolan Ren, Caixia Dong. **Maiji CDC:** Hui Zhang, Enke Mao, Xiaoping Wang, Tao Wang, Xi Zhang. **Henan Provincial CDC:** Kai Kang, Shixian Feng, Huizi Tian, Lei Fan. **Huixian CDC:** XiaoLin Li, Huarong Sun, Pan He, Xukui Zhang. **Zhejiang Provincial CDC:** Min Yu, Ruying Hu, Hao Wang. **Tongxiang CDC:**

Xiaoyi Zhang, Yuan Cao, Kaixu Xie, Lingli Chen, Dun Shen. **Hunan Provincial CDC:** Xiaojun Li, Donghui Jin, Li Yin, Huilin Liu, Zhongxi Fu. **Liuyang CDC:** Xin Xu, Hao Zhang, Jianwei Chen, Yuan Peng, Libo Zhang, Chan Qu.

#### Supplementary Note 1: Array Design

The China Kadoorie Biobank includes participants genotyped on two custom Affymetrix Axiom arrays: CKB1 and CKB2. These platforms differ in probe content and sample ascertainment: CKB1 was used primarily in nested case-control studies and oversampled individuals with major cardiovascular and respiratory disease, while CKB2 was applied to a population-representative sample set with improved probe density [1]. To minimise platform effects, all association analyses were restricted to probes present on both arrays (the shared probe set), and array version was included as a covariate in all regression models alongside study region and ancestry principal components. CNV calling was performed separately within each region–array combination, yielding 20 independent subpopulations subsequently combined for association testing.

##### Empirical validation of pooled analysis at reported loci

To assess whether pooling across array versions distorted inference at reported loci, we performed targeted array-version-stratified validation at the lead probe for each genome-wide significant CNVR–phenotype association (Figures S3–S5). For each lead probe, we evaluated: (i) array-version-specific carrier counts and probe-level masking exclusions; (ii) array-version-stratified effect estimates fitted using the same covariates as the primary RINT analysis; and (iii) a formal CNV$\times$array version interaction test.

**Carrier counts and probe-level masking.** All reported loci were supported by non-zero carriers in both array versions, confirming that no association was attributable to a single array version. Probe-level exclusions due to opposite-type masking were modest (median excluded: 5 in CKB1 and 3 in CKB2), and no masking occurred in mirror-model analyses. As expected from the larger sample size of CKB2, carrier counts were consistently higher in CKB2 than in CKB1 (median carriers: 10 in CKB1 vs 36 in CKB2), with the sparsest array-version-stratified estimates occurring at loci with very low CKB1 carrier counts (minimum = 5).

**Array-version-stratified effect estimates.** Effect estimates were directionally consistent across array versions for all 26 reported lead probe–phenotype associations, with no sign discordance observed. The largest deviations between array-version-specific estimates were concentrated among loci with very low CKB1 carrier counts (Figures S4 and S5). Notably, these visually discordant points were dominated by adiposity-associated duplication signals at 16p11.2, a recurrent CNV locus with well-established dosage effects in the literature. We therefore interpret these deviations as sparse-data instability in the smaller array version rather than biologically meaningful array-version-specific heterogeneity.

**CNV×array version interaction tests.** Interaction tests were initially performed on 27 candidate signals, comprising the 26 reported lead probe–phenotype associations plus the 12p12.1 mirror-model signal; the latter was subsequently excluded from reported results as it had zero deletion carriers at the lead probe and was entirely duplication-driven (see Results). Of these 27, 6 reached nominal significance ($P < 0.05$), and none survived Bonferroni correction for 26 tests after excluding the 12p12.1 mirror signal (threshold $P < 0.0019$; Figure S5). All nominal interaction signals were confined to loci with very low CKB1 carrier counts (5–7 carriers), where array-version-stratified estimates are inherently unstable and associated with wide confidence intervals. Notably, all nominal interaction signals arose from duplication models at the 16p11.2 locus, a well-established recurrent CNV region with large, well-replicated dosage effects in anthropometric traits. The most extreme interaction signal (weight, 16p11.2 duplication, $P = 0.0056$) occurred at a locus with only 5 CKB1 carriers. More generally, the most extreme interaction statistics were concentrated at loci with the lowest CKB1 carrier counts, consistent with sparse-data instability rather than true array-version-specific heterogeneity. A meta-analytic approach would be unlikely to alter this conclusion: with only a handful of carriers contributing minimal inverse-variance weight, meta-analysis and pooled regression are functionally similar, whereas pooled regression is more stable and avoids estimating between-array-version heterogeneity from sparse data.

Taken together, these analyses demonstrate that pooled associations were not driven by a single array version and that covariate adjustment for array version adequately controlled for platform differences. The main apparent outliers arose at loci with extremely low CKB1 carrier counts, particularly at the well-established 16p11.2 adiposity locus, supporting the interpretation that these reflect unstable array-version-specific estimation rather than failure of the pooled analysis.

**Figure S1. Study flowchart**

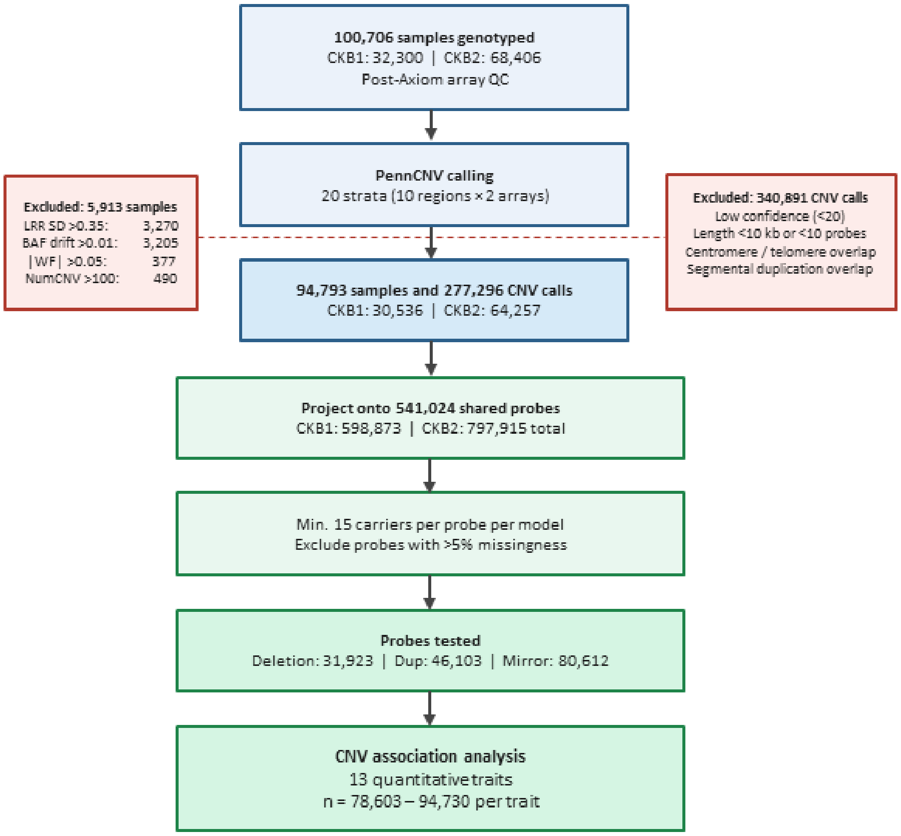


Flowchart summarising sample and CNV quality control and the association analysis pipeline. Of 105,408 samples genotyped (33,408 on CKB1; 72,000 on CKB2), 100,706 passed upstream genotyping quality control (32,300 on CKB1; 68,406 on CKB2) as detailed in Walters et al.[1], and served as input to the CNV calling pipeline described here. Left branch: sample-level quality control applied to these 100,706 individuals. Right branch: CNV calling performed separately for each of 20 region–array strata using PennCNV, followed by call-level quality control. Both branches converge at projection onto the 541,024 probes shared between array versions, after which probe-level carrier thresholds and missingness filters were applied prior to association testing.

**Figure S2. Distribution of CNV calling quality control metrics across CKB genotyped population**

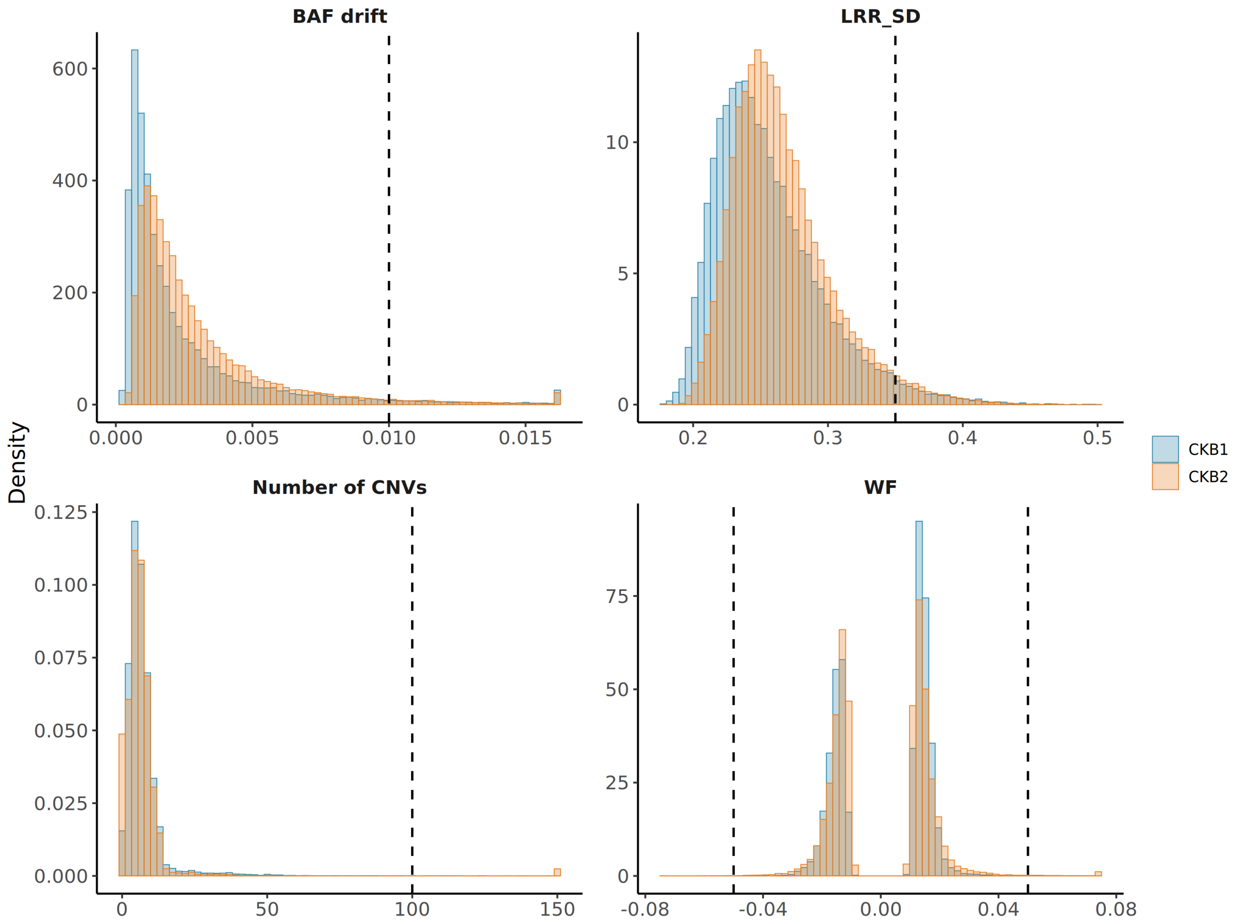


Distributions are stratified by genotyping array version (CKB1, pink; CKB2, teal). Dashed lines indicate exclusion thresholds: BAF drift > 0.01, LRR SD > 0.35, number of CNVs > 100, and |WF| > 0.05. Failure rates by metric were: LRR SD (CKB1 3.1%, CKB2 3.3%), BAF drift (CKB1 3.2%, CKB2 3.2%), WF (CKB1 <0.1%, CKB2 0.5%), and number of CNVs (CKB1 <0.1%, CKB2 0.7%). After joint application of all filters, 30,536 of 32,265 CKB1 samples (94.6%) and 64,257 of 68,441 CKB2 samples (93.9%) were retained, leaving 94,793 individuals (94.1%) for association analyses, of whom 94,730 had phenotypic data available and contributed to association analyses.

#### Figure S3. Array-version-stratified and pooled effect estimates at each lead probe–phenotype association

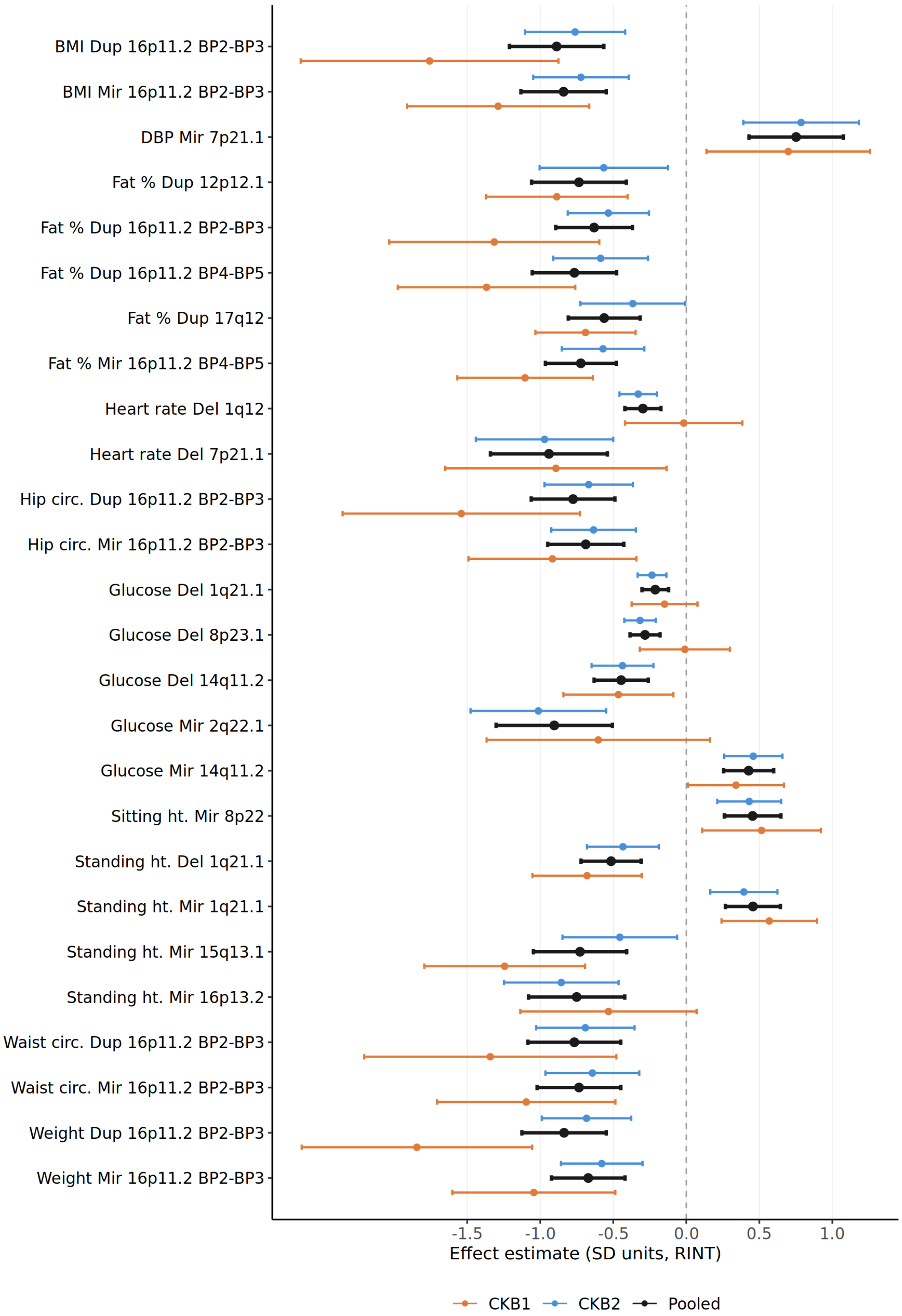


Effect estimates are on the RINT scale. Orange = CKB1, blue = CKB2, black = pooled; points are effect estimates and bars are 95% confidence intervals.

**Figure S4. Lead probe carrier counts at each reported association, stratified by array version**

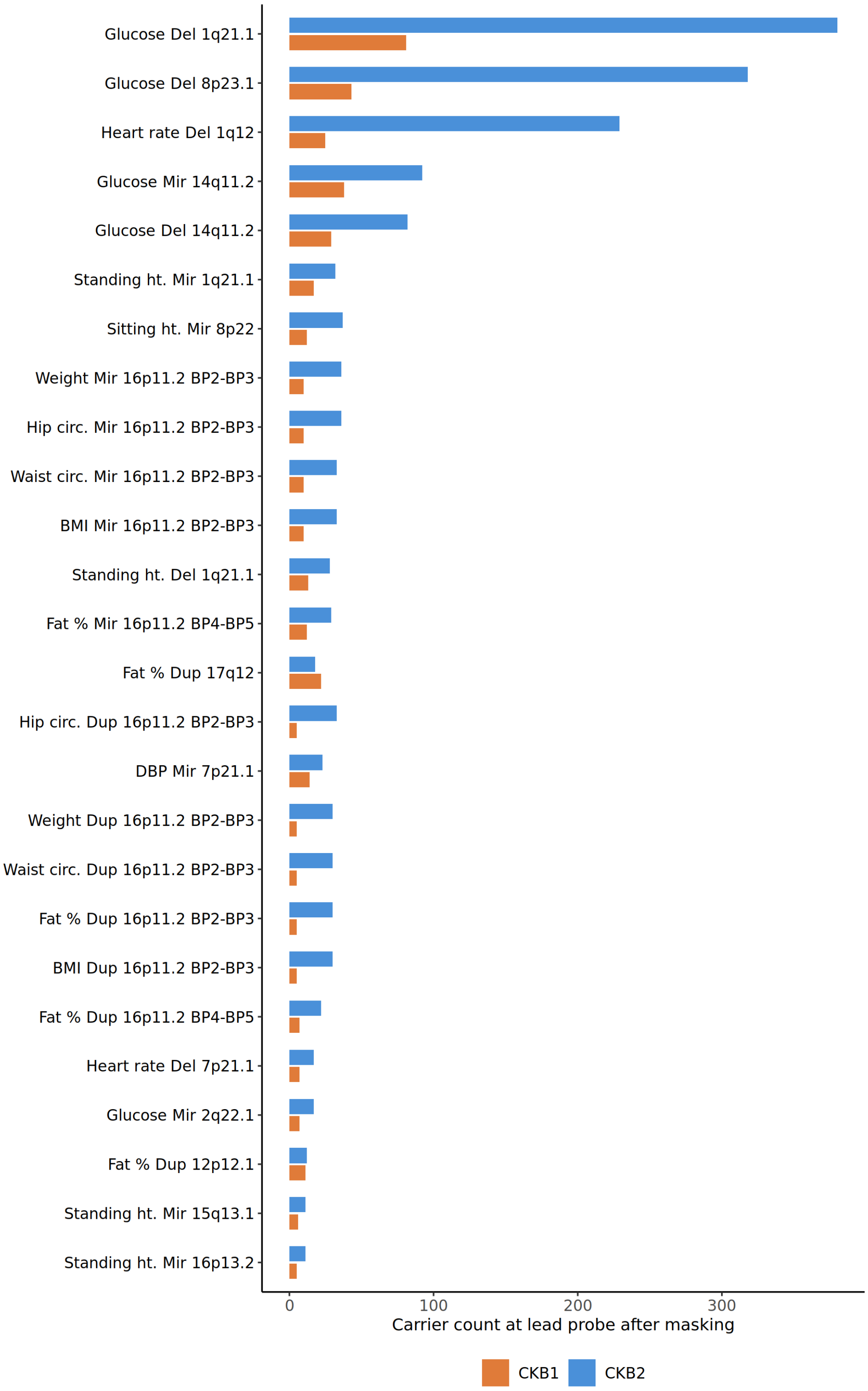


Counts are shown after probe-level masking for each of the 26 reported lead probe–phenotype associations. Orange = CKB1, blue = CKB2.

**Figure S5. CNV×array version interaction significance versus CKB1 carrier count at the lead probe**

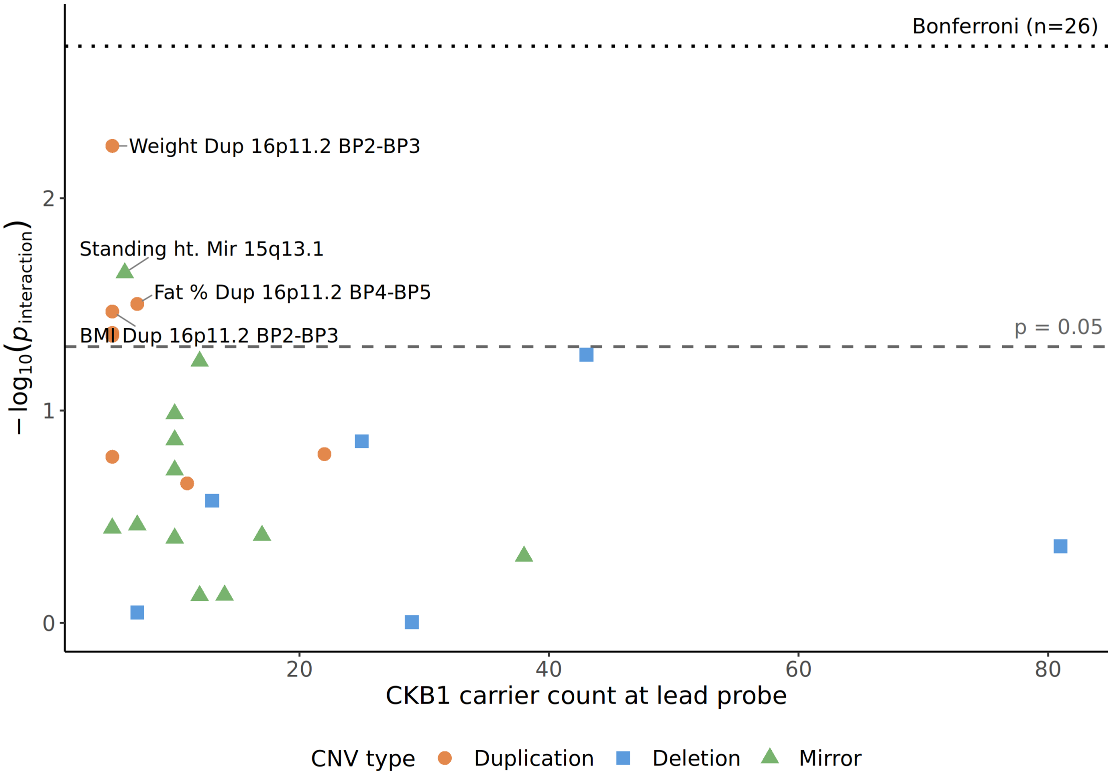


Dashed line = nominal $P = 0.05$; dotted line = Bonferroni threshold ($P = 0.0019$, $n = 26$ tests).

**Figure S6. Unadjusted CNV carrier group distributions for BMI at 16p11.2 BP2–BP3**

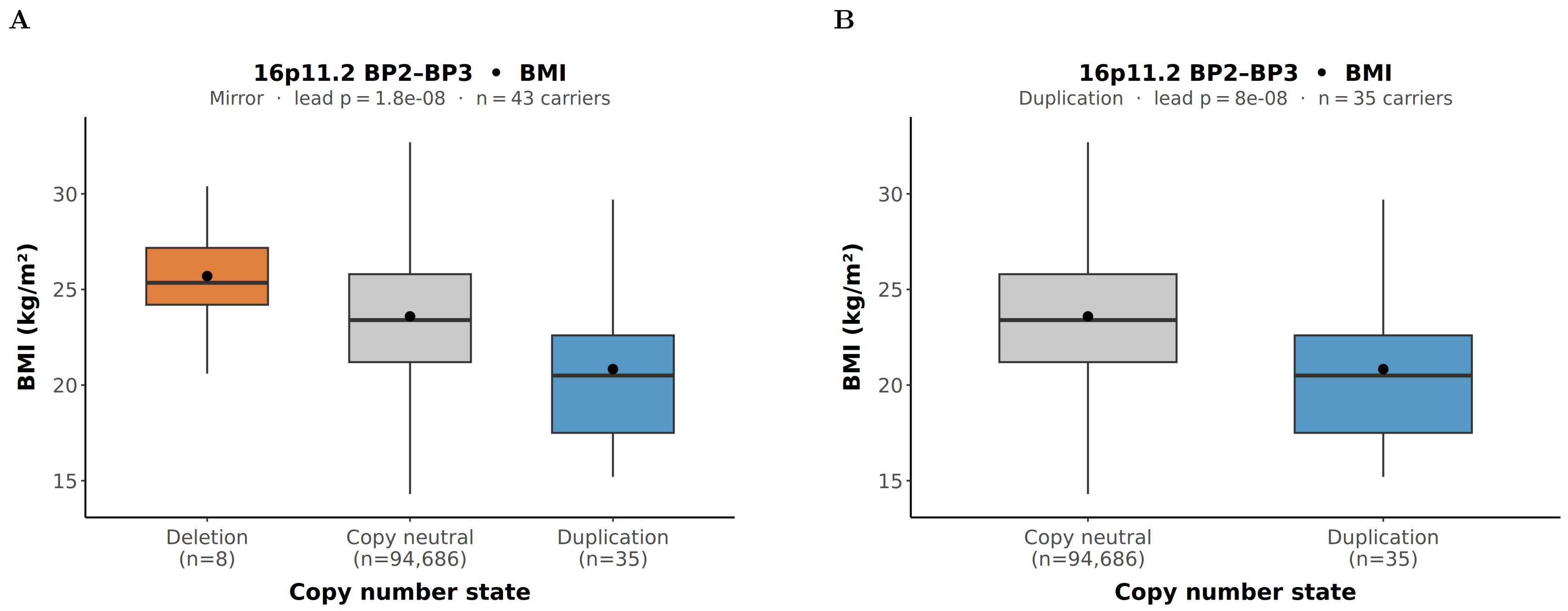


**(A)** Mirror model: mean (median) BMI was 25.7 (25.4) kg/m$^2$ in deletion carriers (n = 8), 23.7 (23.4) kg/m$^2$ in copy-neutral individuals (n = 94,686), and 20.8 (20.5) kg/m$^2$ in duplication carriers (n = 35). **(B)** Duplication model: mean (median) BMI was 20.8 (20.5) kg/m$^2$ in duplication carriers (n = 35) and 23.7 (23.4) kg/m$^2$ in copy-neutral individuals (n = 94,686). Mirror and duplication-model signals represent the same locus. Boxplots show unadjusted phenotype distributions; whiskers extend to 1.5×IQR and filled circles indicate group means. Association p-values are from linear regression adjusting for age, age$^2$, sex, region, array type, and the first 11 genetic principal components.

**Figure S7. Unadjusted CNV carrier group distributions for diastolic blood pressure at 7p21.1**

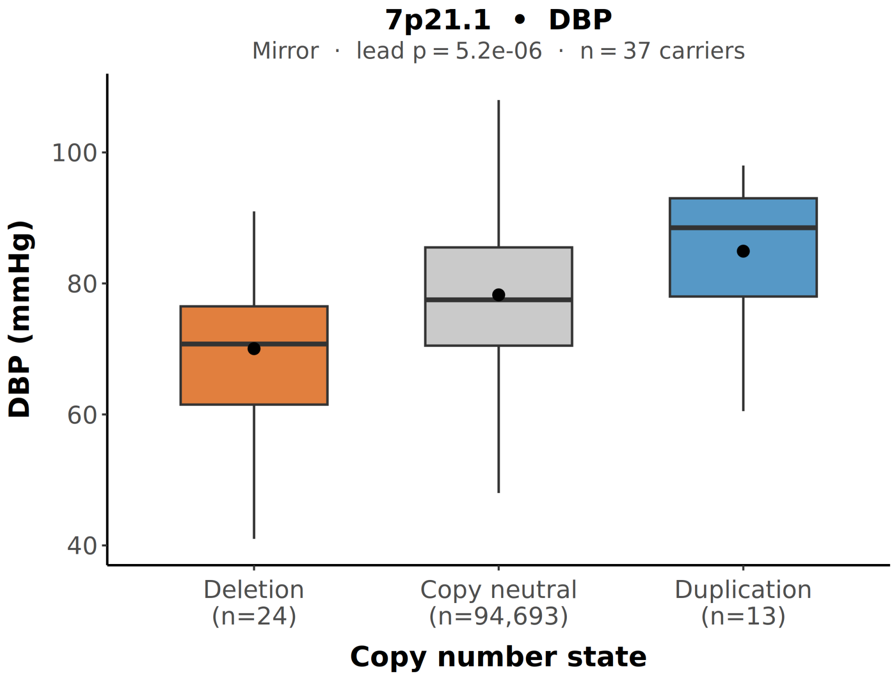


Mirror model: mean (median) DBP was 70.0 (70.8) mmHg in deletion carriers (n = 24), 78.6 (77.5) mmHg in copy-neutral individuals (n = 94,693), and 84.9 (88.5) mmHg in duplication carriers (n = 13).

**Figure S8. Unadjusted CNV carrier group distributions for body fat percentage at 12p12.1**

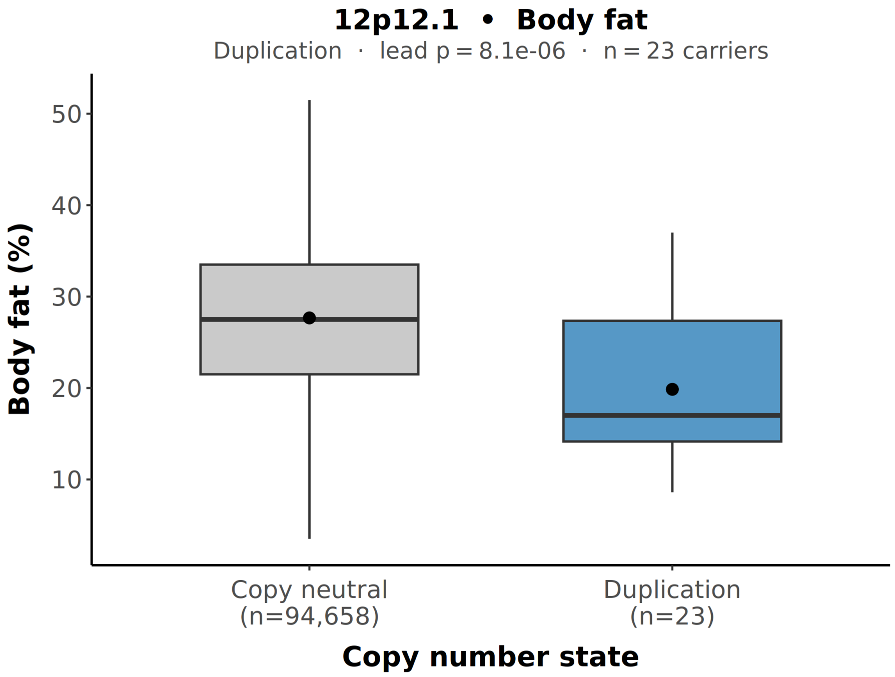


Duplication model: mean (median) body fat was 19.9% (17.0%) in duplication carriers (n = 23) and 27.7% (27.5%) in copy-neutral individuals (n = 94,658). A mirror-model signal was also identified at this locus but had zero deletion carriers at the lead probe and is not shown separately.

**Figure S9. Unadjusted CNV carrier group distributions for body fat percentage at 16p11.2 BP4–BP5**

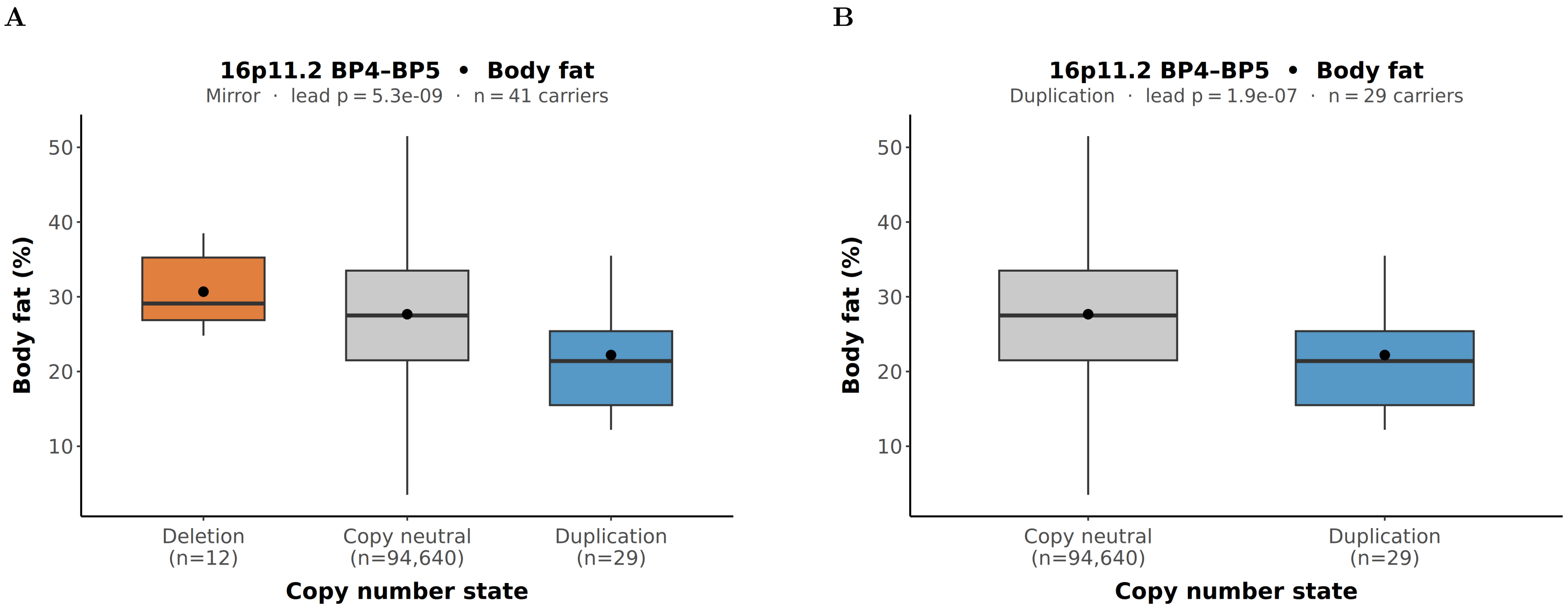


**(A)** Mirror model: mean (median) body fat was 30.7% (29.1%) in deletion carriers (n = 12), 27.7% (27.5%) in copy-neutral individuals (n = 94,640), and 22.2% (21.4%) in duplication carriers (n = 29). **(B)** Duplication model: mean (median) body fat was 22.2% (21.4%) in duplication carriers (n = 29) and 27.7% (27.5%) in copy-neutral individuals (n = 94,640). Mirror and duplication-model signals represent the same locus.

**Figure S10. Unadjusted CNV carrier group distributions for body fat percentage at 16p11.2 BP2–BP3**

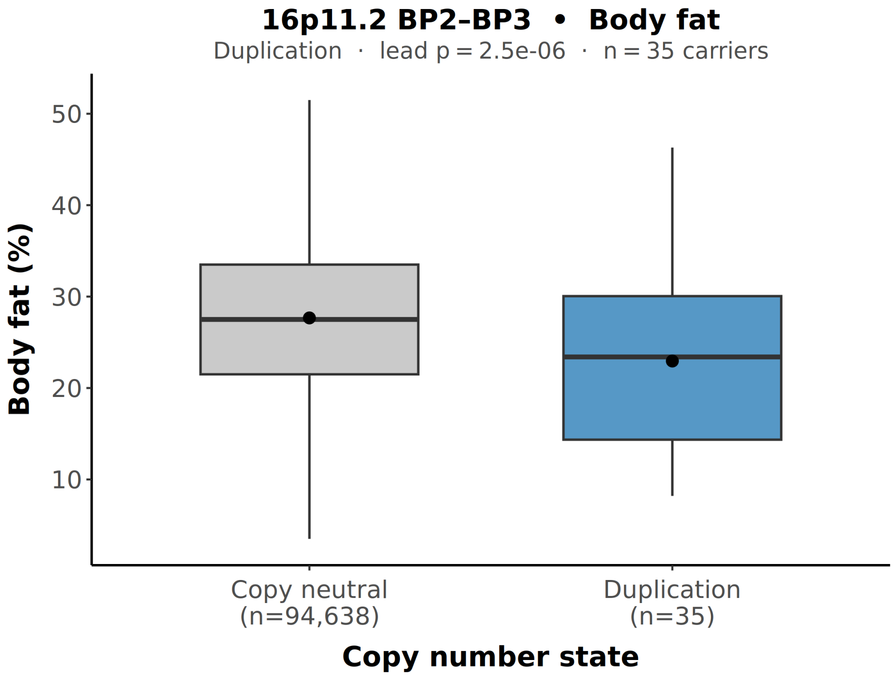


Duplication model: mean (median) body fat was 23.0% (23.4%) in duplication carriers (n = 35) and 27.7% (27.5%) in copy-neutral individuals (n = 94,638). A mirror-model signal was also identified at this locus but had zero deletion carriers at the lead probe and is not shown separately.

**Figure S11. Unadjusted CNV carrier group distributions for body fat percentage at 17q12**

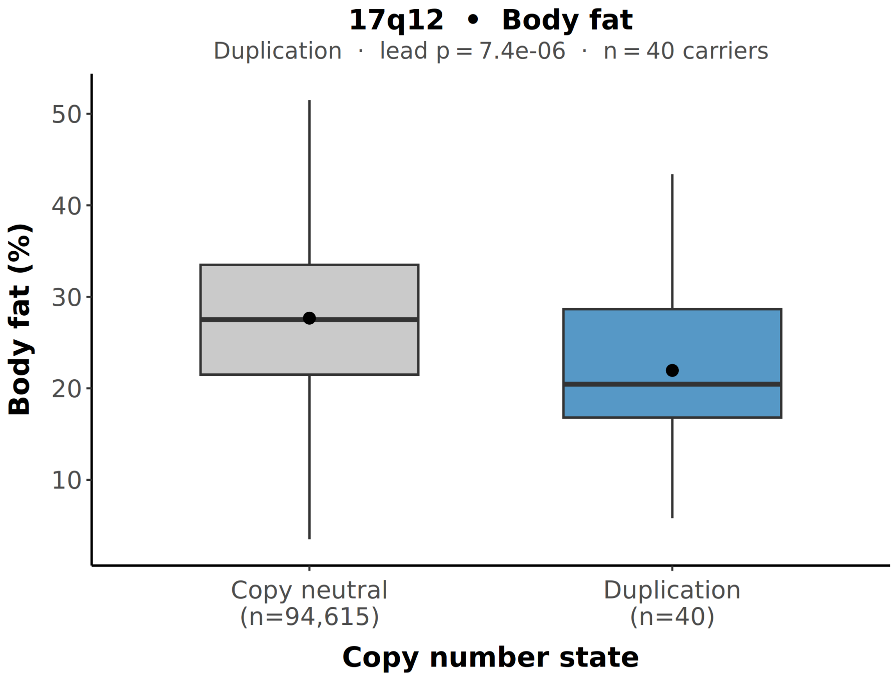


Duplication model: mean (median) body fat was 22.0% (20.5%) in duplication carriers (n = 40) and 27.7% (27.5%) in copy-neutral individuals (n = 94,615).

**Figure S12. Unadjusted CNV carrier group distributions for resting heart rate at 1q12**

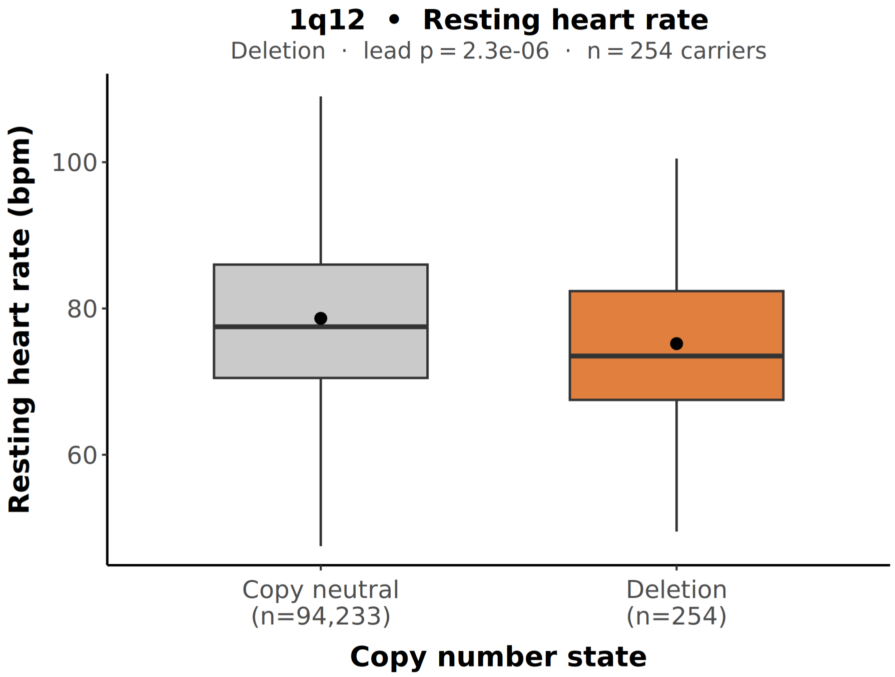


Deletion model: mean (median) heart rate was 75.2 (73.5) bpm in deletion carriers (n = 254) and 79.1 (78.0) bpm in copy-neutral individuals (n = 94,233).

**Figure S13. Unadjusted CNV carrier group distributions for resting heart rate at 7p21.1**

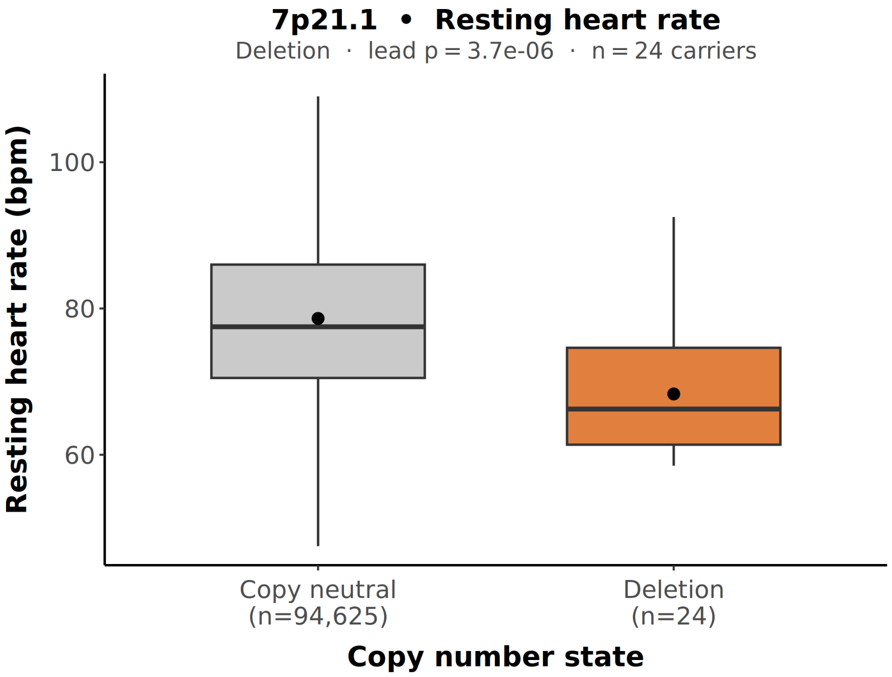


Deletion model: mean (median) heart rate was 68.3 (66.3) bpm in deletion carriers (n = 24) and 79.1 (78.0) bpm in copy-neutral individuals (n = 94,625).

**Figure S14. Unadjusted CNV carrier group distributions for hip circumference at 16p11.2 BP2–BP3**

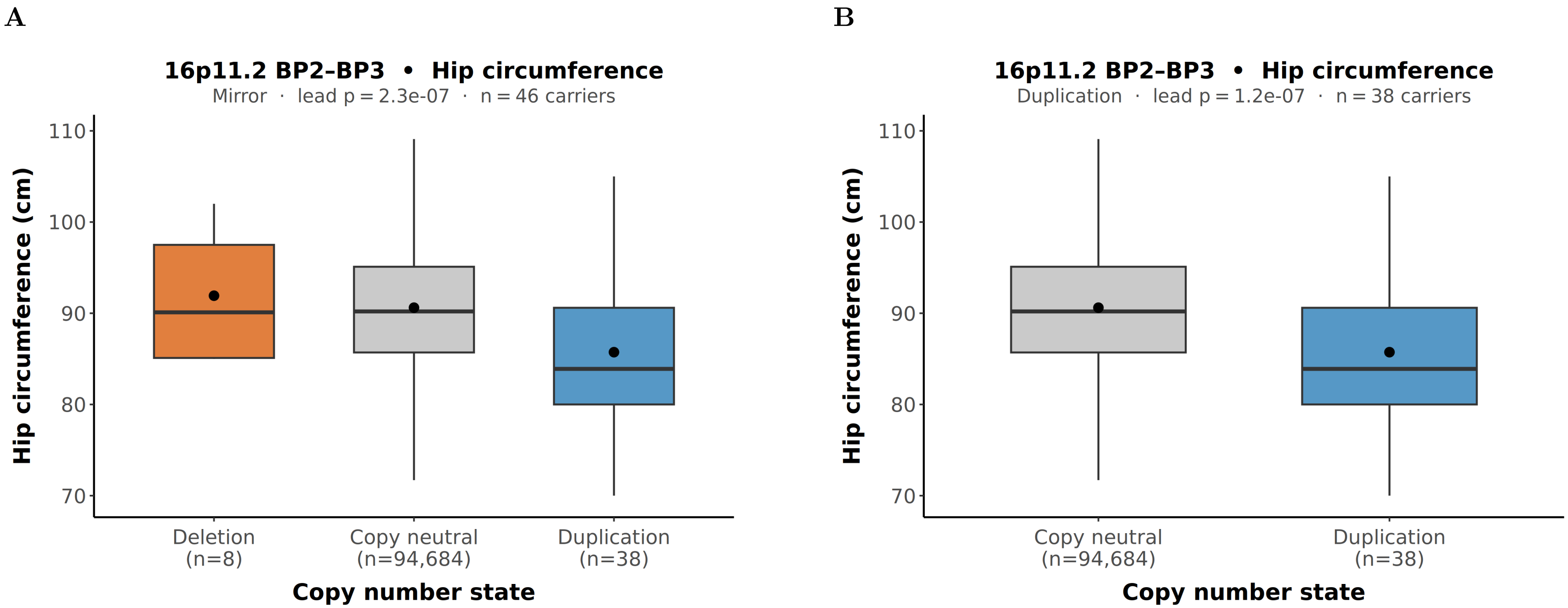


**(A)** Mirror model: mean (median) hip circumference was 91.9 (90.1),cm in deletion carriers (n,=,8), 90.8 (90.3),cm in copy-neutral individuals (n,=,94,684), and 85.7 (83.9),cm in duplication carriers (n,=,38). **(B)** Duplication model: mean (median) hip circumference was 85.7 (83.9),cm in duplication carriers (n,=,38) and 90.8 (90.3),cm in copy-neutral individuals (n,=,94,684). Mirror and duplication-model signals represent the same locus.

**Figure S15. Unadjusted CNV carrier group distributions for random glucose at 1q21.1**

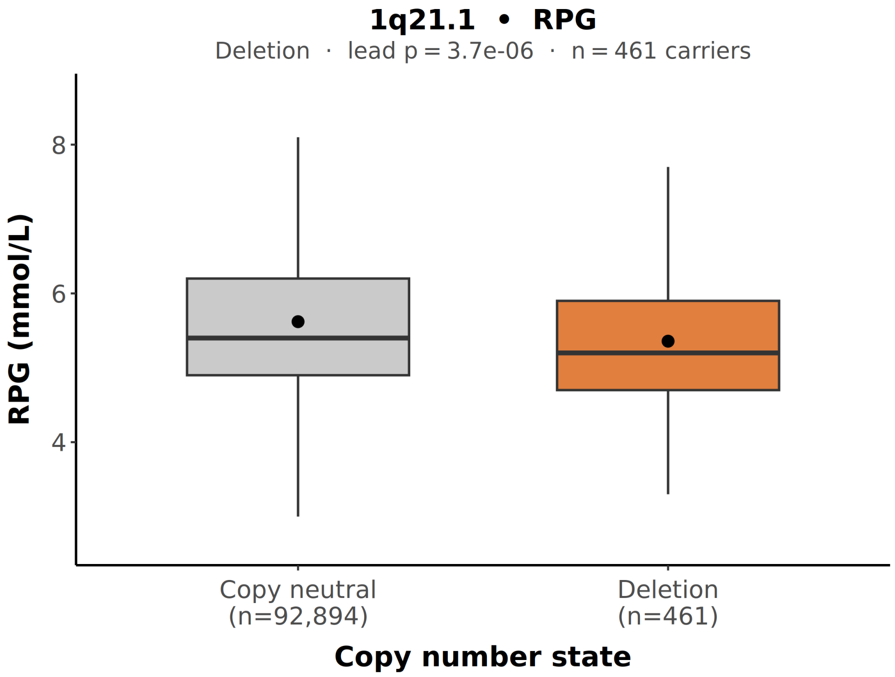


Deletion model: mean (median) random glucose was 5.77 (5.3) mmol/L in deletion carriers (n = 461) and 6.11 (5.6) mmol/L in copy-neutral individuals (n = 92,894).

**Figure S16. Unadjusted CNV carrier group distributions for random glucose at 2q22.1**

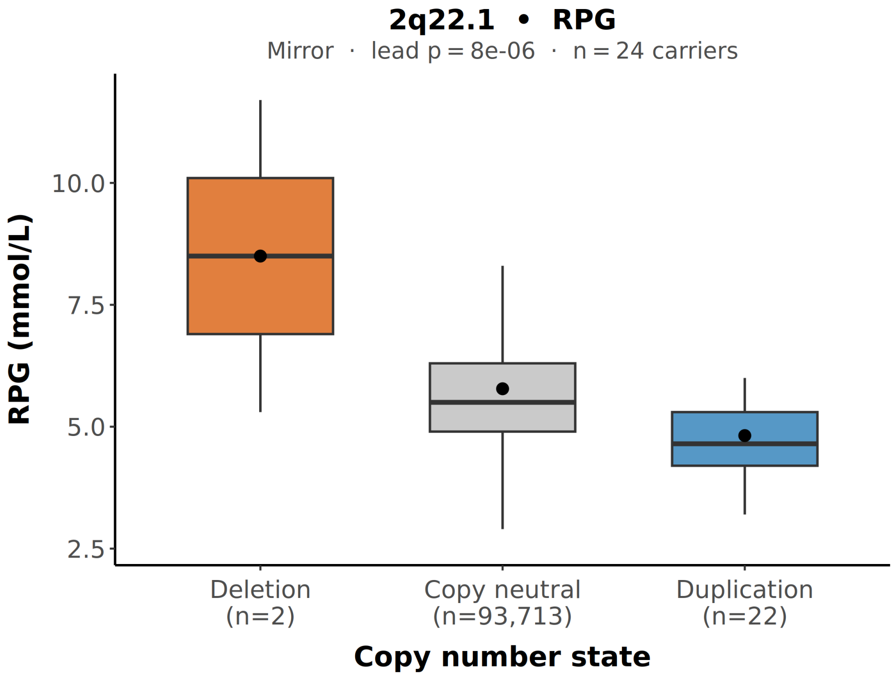


Mirror model: mean (median) random glucose was 8.5 (8.5) mmol/L in deletion carriers (n = 2), 6.11 (5.6) mmol/L in copy-neutral individuals (n = 93,713), and 4.82 (4.65) mmol/L in duplication carriers (n = 22). Note: only 2 deletion carriers were present at the lead probe; this signal should be interpreted with caution.

**Figure S17. Unadjusted CNV carrier group distributions for random glucose at 8p23.1**

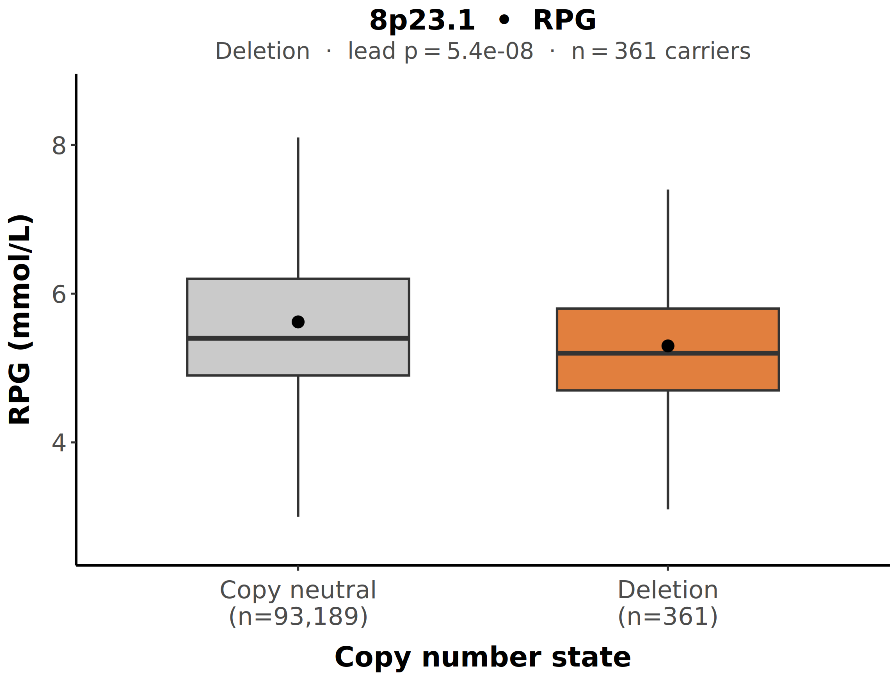


Deletion model: mean (median) random glucose was 5.69 (5.2) mmol/L in deletion carriers (n = 361) and 6.11 (5.6) mmol/L in copy-neutral individuals (n = 93,189).

**Figure S18. Unadjusted CNV carrier group distributions for random glucose at 14q11.2**

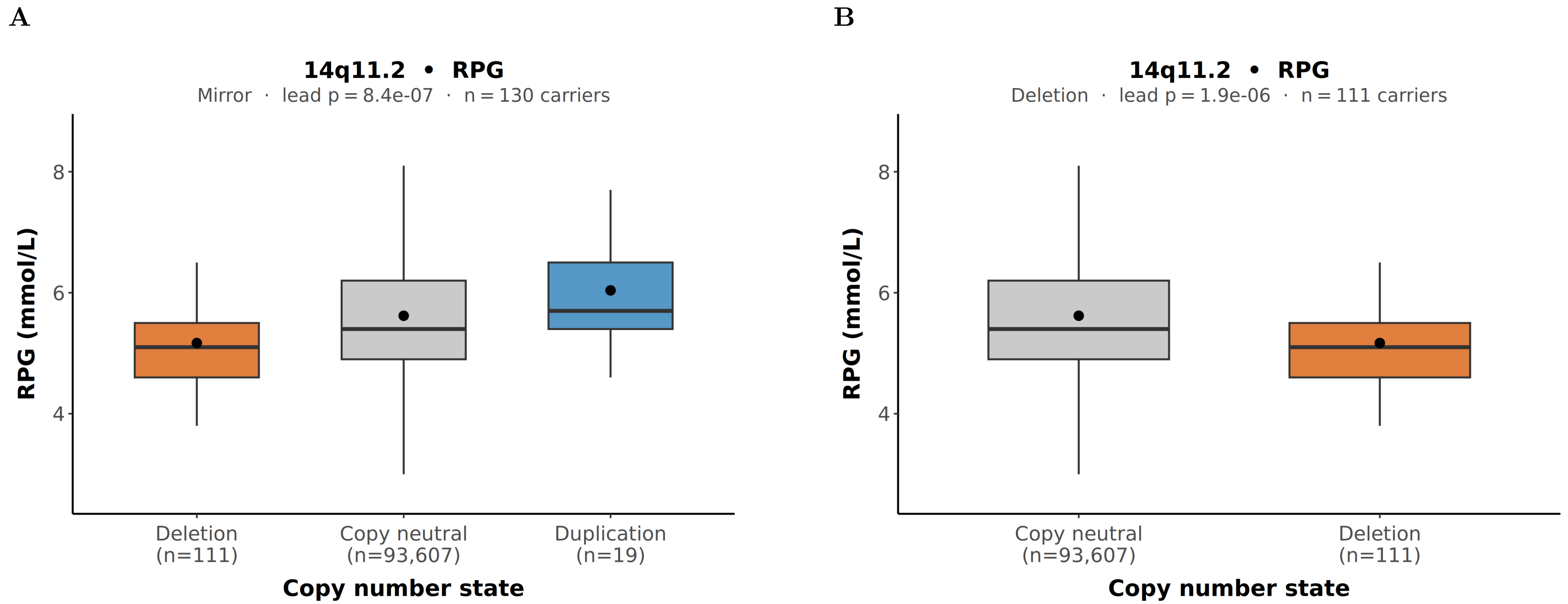


**(A)** Mirror model: mean (median) random glucose was 5.55 (5.1) mmol/L in deletion carriers (n = 111), 6.11 (5.6) mmol/L in copy-neutral individuals (n = 93,607), and 6.23 (5.7) mmol/L in duplication carriers (n = 19). **(B)** Deletion model: mean (median) random glucose was 5.55 (5.1) mmol/L in deletion carriers (n = 111) and 6.11 (5.6) mmol/L in copy-neutral individuals (n = 93,607). Mirror and deletion-model signals represent the same locus.

**Figure S19. Unadjusted CNV carrier group distributions for sitting height at 8p22**

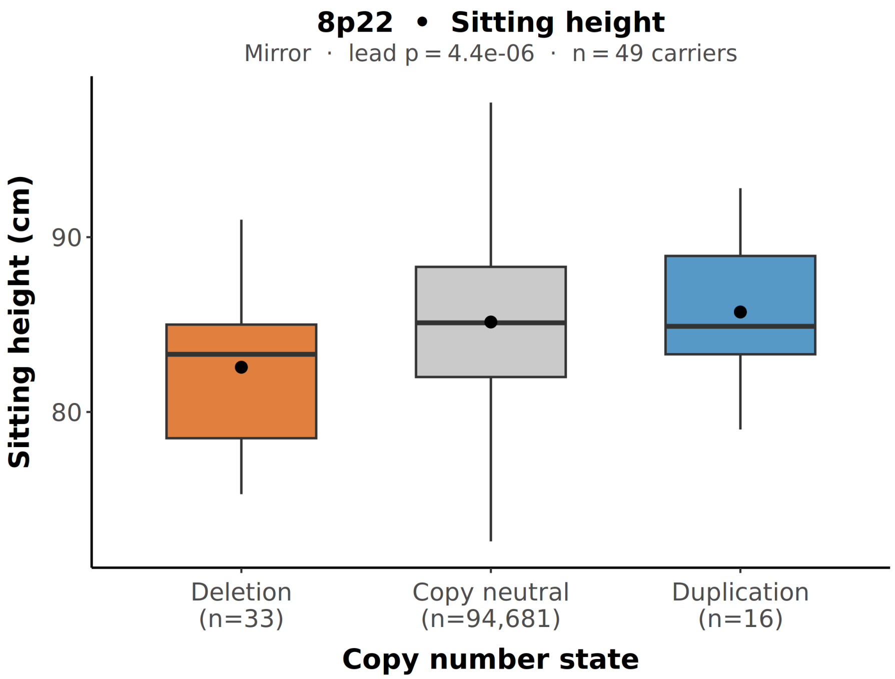


Mirror model: mean (median) sitting height was 82.6 (83.3),cm in deletion carriers (n,=,33), 85.1 (85.1),cm in copy-neutral individuals (n,=,94,681), and 85.7 (84.9),cm in duplication carriers (n,=,16).

**Figure S20. Unadjusted CNV carrier group distributions for standing height at 1q21.1**

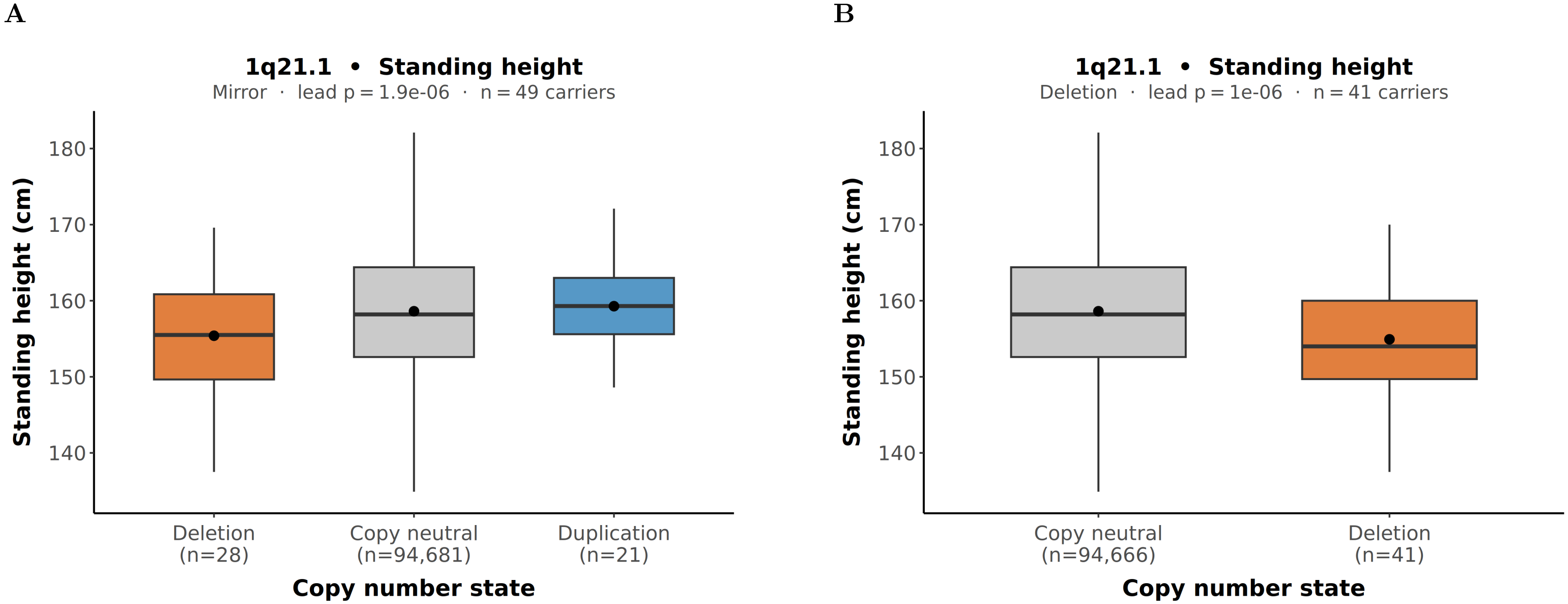


**(A)** Mirror model: mean (median) standing height was 155.4 (155.5),cm in deletion carriers (n,=,28), 158.6 (158.2),cm in copy-neutral individuals (n,=,94,681), and 159.3 (159.3),cm in duplication carriers (n,=,21). **(B)** Deletion model: mean (median) standing height was 154.9 (154.0),cm in deletion carriers (n,=,41) and 158.6 (158.2),cm in copy-neutral individuals (n,=,94,666). Mirror and deletion-model signals represent the same locus.

**Figure S21. Unadjusted CNV carrier group distributions for standing height at 15q13.1**

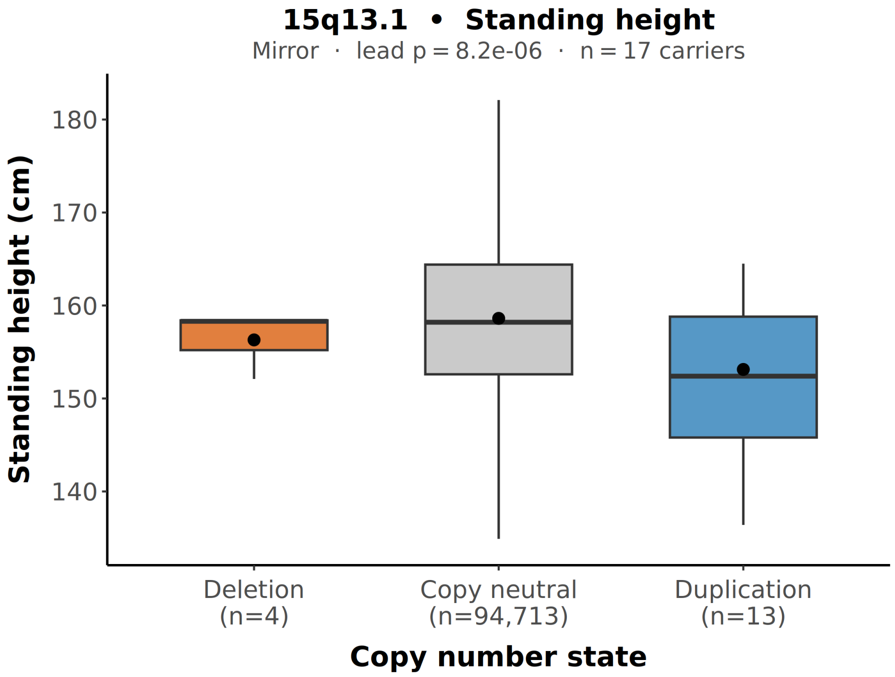


Mirror model: mean (median) standing height was 1638.5 (1584) mm in deletion carriers (n = 4), 1586.2 (1582) mm in copy-neutral individuals (n = 94,713), and 1531.2 (1524) mm in duplication carriers (n = 13). Note: only 4 deletion carriers were present at the lead probe; this signal should be interpreted with caution.

**Figure S22. Unadjusted CNV carrier group distributions for standing height at 16p13.2**

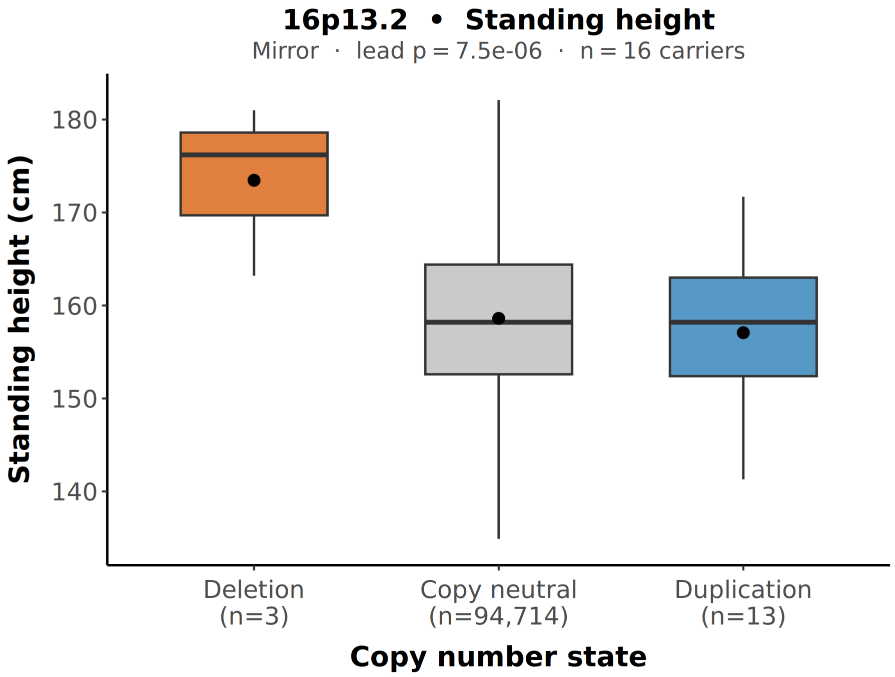


Mirror model: mean (median) standing height was 163.9 (158.4),cm in deletion carriers (n,=,4), 158.6 (158.2),cm in copy-neutral individuals (n,=,94,713), and 153.1 (152.4),cm in duplication carriers (n,=,13).

**Figure S23. Unadjusted CNV carrier group distributions for waist circumference at 16p11.2 BP2–BP3**

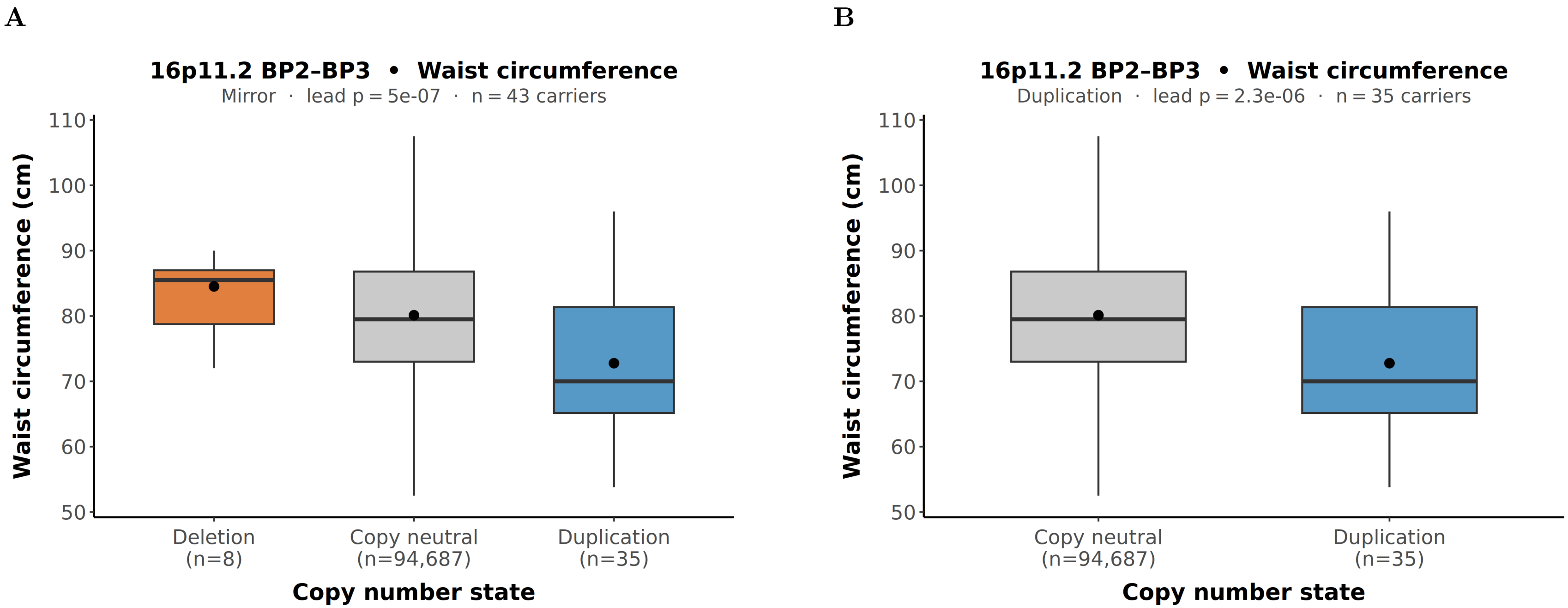


**(A)** Mirror model: mean (median) waist circumference was 84.5 (85.5),cm in deletion carriers (n,=,8), 80.2 (79.6),cm in copy-neutral individuals (n,=,94,687), and 72.8 (70.0),cm in duplication carriers (n,=,35). **(B)** Duplication model: mean (median) waist circumference was 72.8 (70.0),cm in duplication carriers (n,=,35) and 80.2 (79.6),cm in copy-neutral individuals (n,=,94,687). Mirror and duplication-model signals represent the same locus.

**Figure S24. Unadjusted CNV carrier group distributions for weight at 16p11.2 BP2–BP3**

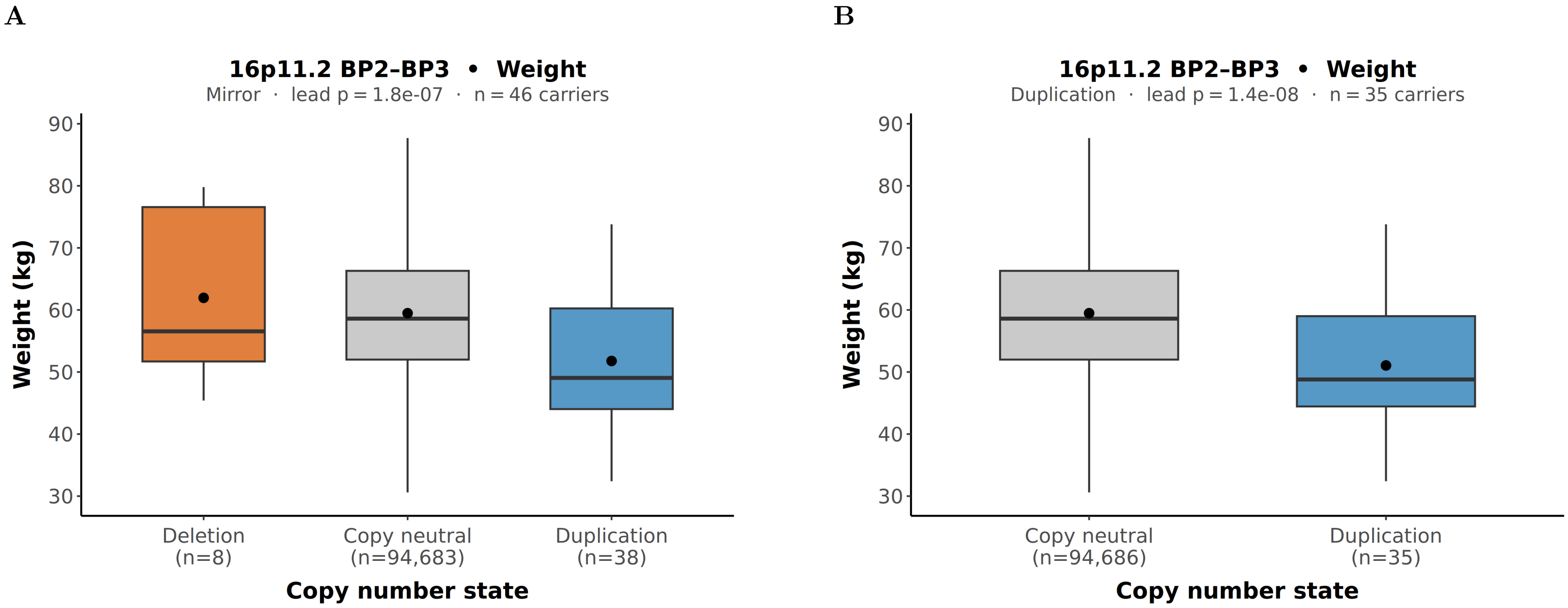


**(A)** Mirror model: mean (median) weight was 61.9 (56.6) kg in deletion carriers (n = 8), 59.7 (58.7) kg in copy-neutral individuals (n = 94,683), and 51.8 (49.1) kg in duplication carriers (n = 38). **(B)** Duplication model: mean (median) weight was 51.1 (48.8) kg in duplication carriers (n = 35) and 59.7 (58.7) kg in copy-neutral individuals (n = 94,686). Mirror and duplication-model signals represent the same locus.

**Figure S25. Concordance of effect estimates between the full sample and an unrelated subsample**

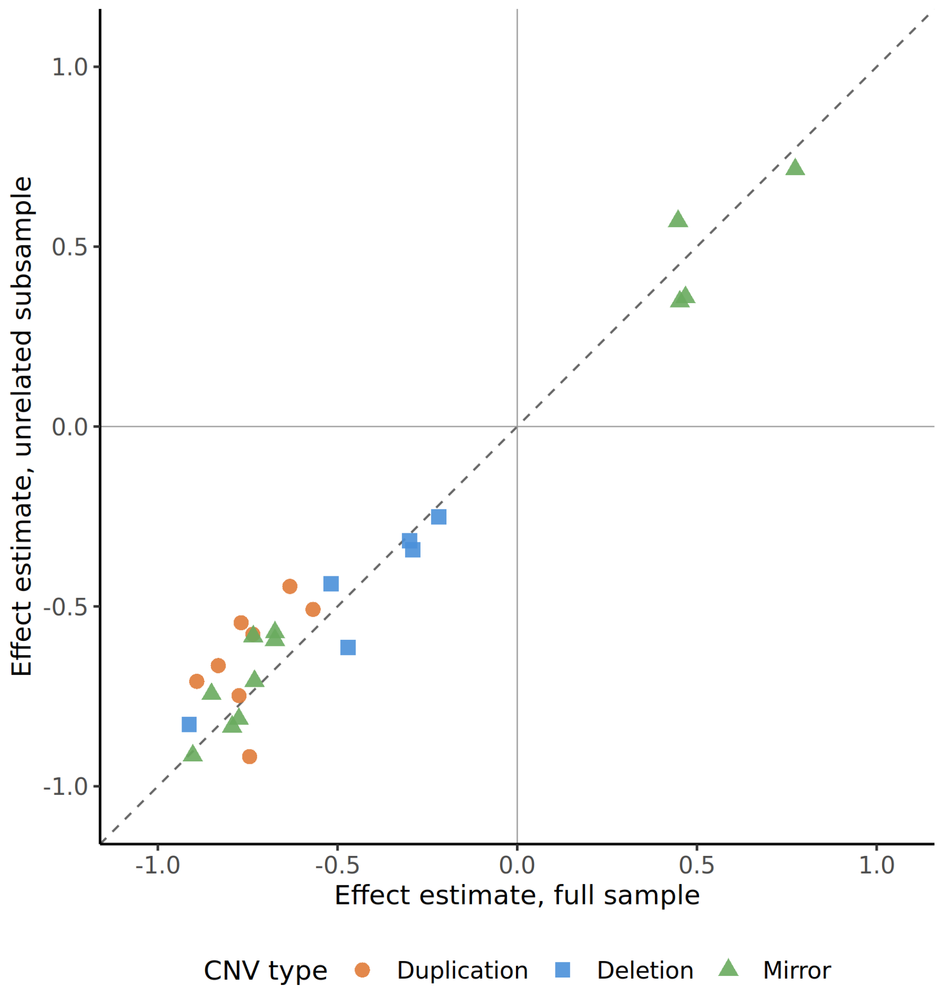


Each point represents one of the 26 reported lead probe–phenotype associations (n = 71,477–72,183 depending on trait), with effect estimates on the inverse rank-normal transformed scale. Dashed line = identity. Colour and shape indicate CNV model: orange circles = duplication, blue squares = deletion, green triangles = mirror. All 26 signals showed directional concordance between the full and unrelated samples.
